# Economic vulnerability and poor service delivery made it more difficult for shack-dwellers to comply with COVID-19 restrictions: The impracticability and inequitable burden of universal/unstratified public health policies

**DOI:** 10.1101/2022.04.07.22273499

**Authors:** GTH Ellison, RB Mattes, H Rhoma, T De Wet

## Abstract

In South Africa, demand for housing close to viable/sustained sources of employment has far outstripped supply; and the size of the population living in temporary structures/shacks (and in poorly serviced informal settlements) has continued to increase. While such dwellings and settlements pose a number of established risks to the health of their residents, the present study aimed to explore whether they might also undermine the potential impact of regulations intended to safeguard public health, such as the stringent lockdown restrictions imposed to curb the spread of COVID-19 in 2020 and 2021. Using a representative sample of 1,381 South African households surveyed in May-June 2021, the present study found that respondents in temporary structures/shacks were more likely to report non-compliance (or difficulty in complying) with lockdown restrictions when compared to those living in traditional/formal houses/flats/rooms/hostels (OR:1.61; 95%CI:1.06-2.45). However, this finding was substantially attenuated and lost precision following adjustment for preceding sociodemographic and economic determinants of housing quality (adjusted OR:1.20; 95%CI:0.78-1.87). Instead, respondents were far more likely to report non-compliance (or difficulty in complying) with COVID-19 lockdown restrictions if their dwellings lacked private/indoor toilet facilities (adjusted OR:1.56; 95%CI:1.08,2.22) or they were ‘Black/African’, young, poorly educated and under-employed (regardless of: their socioeconomic position, or whether they resided in temporary structures/shacks, respectively). Restrictions imposed to safeguard public health need to be more sensitively designed to accommodate the critical role that poverty and inadequate service delivery play in limiting the ability of residents living in temporary structures/shacks and inadequately serviced dwellings/settlements to comply. **[250/250 words]**

**Significance of the main findings:**

- South Africans living in temporary structures/shacks are more likely to: be poorly educated and under-employed; with fewer assets and limited access to basic household services.
- Poverty and inadequate service delivery were more important determinants of compliance with COVID-19 restrictions than housing quality.
- In the absence of improvements in economic circumstances and the delivery of basic household services, restrictions imposed to safeguard public health need to be more sensitively designed to take account of the structural barriers to compliance experienced by households where poverty and/or inadequate service delivery limit their ability to: stay at home; maintain hygiene; and/or practice social distancing. **[100/100 words]**

*“This idea of the “humbling pandemic”*^1^ *does not hold for people whose lives depend on informal economy and movement in the face of heavy restrictions on their respective activities such as… street hustle and domestic work. Therefore, the pandemic response – which employs tactics that come to determine how lives are to be lived – can be seen as an exacerbator of inequalities, by the hands of which precarious circumstances of living are a larger threat than the risk of infection”*^2^
Stefan Ogedengbe (2021: 94)^3^

## Introduction

It is important not to overlook the role that temporary structures/shacks can play in accommodating the needs, aspirations and agency of citizens when public policies fail to provide affordable housing.^4-8^ Nor should we dismiss the role that shared adversity can play in the formation of grassroots social and political movements capable of delivering tangible benefits to the communities involved.^9-12^ Yet while necessity might well be the ‘mother of invention’, there can be little doubt that such dwellings pose multiple challenges to the health and wellbeing of their residents;^13-14^ and that disadvantage, poverty and despair are what more commonly lie behind the “necessity” to take shelter in (or, indeed, to *make* shelters from) temporary structures that: offer inadequate and substandard accommodation; limited protection from the elements, and are at increased risk of catastrophic events (such as fires, floods and storms); while providing little security for their residents and their possessions.^15^ Unsurprisingly, researchers who have explored the many substantive and subtle, direct and indirect contributions that housing can make to health^16^ are scathing in their assessments of temporary structures/shacks, pointing out that the *“physical and socio-economic conditions found in informal settlements are generally hazardous to health and tend to exacerbate the severe socio-economic conditions of the urban poor”*^14^ and contain “*all* [of] *the conditions* [required] *for* [the] *rapid spread* [of infectious disease]: *very high population density, scant access to water and sanitation, widespread poverty and inadequate health infrastructure…”*^10^

For these reasons, the predominant focus of research into the health and wellbeing of households living in temporary structures/shacks – both within the backyards of more permanent/formal dwellings^7,8,17^ and in emerging/established ‘informal settlements’^18,19^ – has been on their social, political and structural determinants, correlates and consequences.^7,13^ Such research has had an important role to play in documenting the scale of the problems facing rapidly urbanising populations, and where internal and international migration, population growth, social change, weak governance and limited resources all conspire to create demand for housing that far outstrips supply.^6,20^ In South Africa these challenges also reflect the enduring legacy of apartheid policies that allocated residential rights on the basis of racialised ‘Population Group’ classifications,^21^ and tightly controlled access to formal housing that was close to viable and sustained sources of employment (particularly where these were also close to areas reserved for those classified as ‘White/European’).^20^ The abolition of these ‘influx control’ statutes in 1986^22^ (and the subsequent repeal of the last of the ‘Group Areas Acts’ [No.36 of 1966] in 1990) has both accentuated and accelerated the ongoing depopulation of South Africa’s more rural provinces as people have sought work and better livelihoods in the country’s urban and industrial centres.^23^ At the same time, the end of apartheid also saw a substantial increase in migration from Africa (and beyond), as migrants sought opportunities in one of the continent’s strongest economies.^24^

Despite these seemingly inexorable trends, and the importance afforded the right to housing in South Africa’s 1996 constitution,^25-27^ a raft of successive government policies and commitments to address the need for additional, affordable housing have demonstrably failed to deliver the quantity and quality required to accommodate the shortfall in housing generated by the rapid rate of urbanisation.^8,12,20,23,27-30^ As a result, the proportion of South Africa’s population living in temporary structures/shacks remains high (at ∼15%) and shows no sign of abating; while any short-term benefits of relocating close(r) to sources of employment^23^ (and any associated benefits in terms of the ‘health selection’ of those involved)^31-32^ are likely to dissipate whenever the economy falters or competition for employment makes wages stagnate, or work opportunities dry up. Indeed, the inherent vulnerability of impoverished households living in temporary structures/shacks places them at greater risk of being trapped in a worsening cycle of poverty, leading to a steady decline in the social and material fabric of communities containing large numbers of such dwellings (and particularly those ‘informal settlements’ where temporary structures/shacks predominate).^17^ These add further, *communal* risks to the physical and mental health problems such communities face, particularly wherever: inadequate water and sanitation services facilitate the spread of infectious disease;^17,19^ the absence of mains electricity makes households reliant on less efficient and more dangerous sources of heat and light;^33^ and social unrest and criminality pose tangible threats to the safety and security of individuals and marginalised groups.^34^ Such factors further accentuate the vulnerability of both households and communities, and further undermine their resilience to cope with or mitigate structural and systemic changes beyond their control.^35,36^

Although these challenges and realities have been well-documented and are widely recognised,^23^ they are often framed in ways that ensure they are simply accepted as an inevitable (or at least an intractable) consequence of external forces over which local authorities, governments and nation states have limited influence (such as ‘market forces’ and ‘globalisation’).^10,12,20,27,37^ When overlooked or dismissed in this way it is not surprising that policy makers fail to acknowledge or accommodate the very particular needs of these communities, and resort to imposing policies with which they are ill-equipped or simply unable to comply.^38^ In the process, it is commonplace for policy makers (and commentators) to mistake the inability of shack-dwellers to comply as an unwillingness to conform – whether that be to land ownership statutes, building regulations, health and safety guidance, or the emergency lockdown restrictions imposed following the onset of the COVID-19 pandemic.^23^ For this reason, the aim of the present study was to examine the determinants, correlates and consequences of residence in a temporary structure/shack twelve months into the COVID-19 pandemic, to better understand the role that disadvantage, poverty and inadequate service delivery might play in the ability of residents to comply with COVID-19 lockdown restrictions.

## Methods

### Data collection

The present study used South African data generated during Round 8 of Afrobarometer (AB-R8; https://afrobarometer.org/) – a series of cross-national surveys which began in 1999 and currently covering 34 African countries. The South African arm of the AB-R8 survey was undertaken by Plus 94 Research (PTY) Ltd (https://plus94.co.za/) between 2^nd^ May and 12^th^ June 2021, and involved trained fieldworkers conducting interviews (in-person and in the language chosen by each respondent) with a nationally representative, random, stratified probability sample of 1,600 adult South Africans.^39^ The sampling units/enumeration areas used followed the sampling frame developed for use by Statistics South Africa’s 2011 Population and Housing Census,^40^ stratified by: province; rural/urban locale; and dominant quasi-racial ‘population group’ – with the distribution of these strata updated in line with Statistics South Africa’s 2016 Community Survey.^41^ This involved a total of 400 enumeration areas randomly selected with probabilities proportionate to the sample size. Within each selected area, four households were selected using pre-set walk patterns originating from randomly selected start points; and within each household, one resident adult (aged ≥18yr) was then randomly selected for interview, yielding an overall sample of 1,600 respondents.

### Analytical design

To examine the putative causal relationships between sociodemographic, economic, household and COVID-19 related determinants, correlates and consequences of: residence in a temporary structure/shack; and ease of compliance with COVID-19 lockdown restrictions, we designed our analyses around a hypothesised causal path diagram (in the form of a directed acyclic graph; see Figure 1, below). This diagram sought to apply temporal logic to identify those (sociodemographic, economic, household and COVID-19 related) features considered likely to have preceded one another in a theoretical temporal sequence/cascade of (time-invariant) ‘events’ (such as respondent gender or ‘Population Group’ classification) and ‘crystallised’ (time-variant) characteristics (such as employment status or respondent/household assets), whose position within this sequence will have been determined by the timing of, and the specific items included in, the AB-R8 survey questionnaire (as well as by the contexts and circumstances under which this questionnaire was answered by survey participants).^42^ In the absence of substantive evidence to the contrary, any variables considered likely to have occurred (or ‘crystallised’) before any given exposure variable were assumed to act as potential (or at the very least, ‘candidate’) confounders, since these can be considered *probabilistic* causes of *both* the specified exposure *and* its subsequent (specified) outcome(s). All such confounders require conditioning (through sampling, stratification or – as here – statistical adjustment) to deliver estimates of ‘total causal effects’ in which the risk of confounding bias has been mitigated.^42^

**Figure 1.**
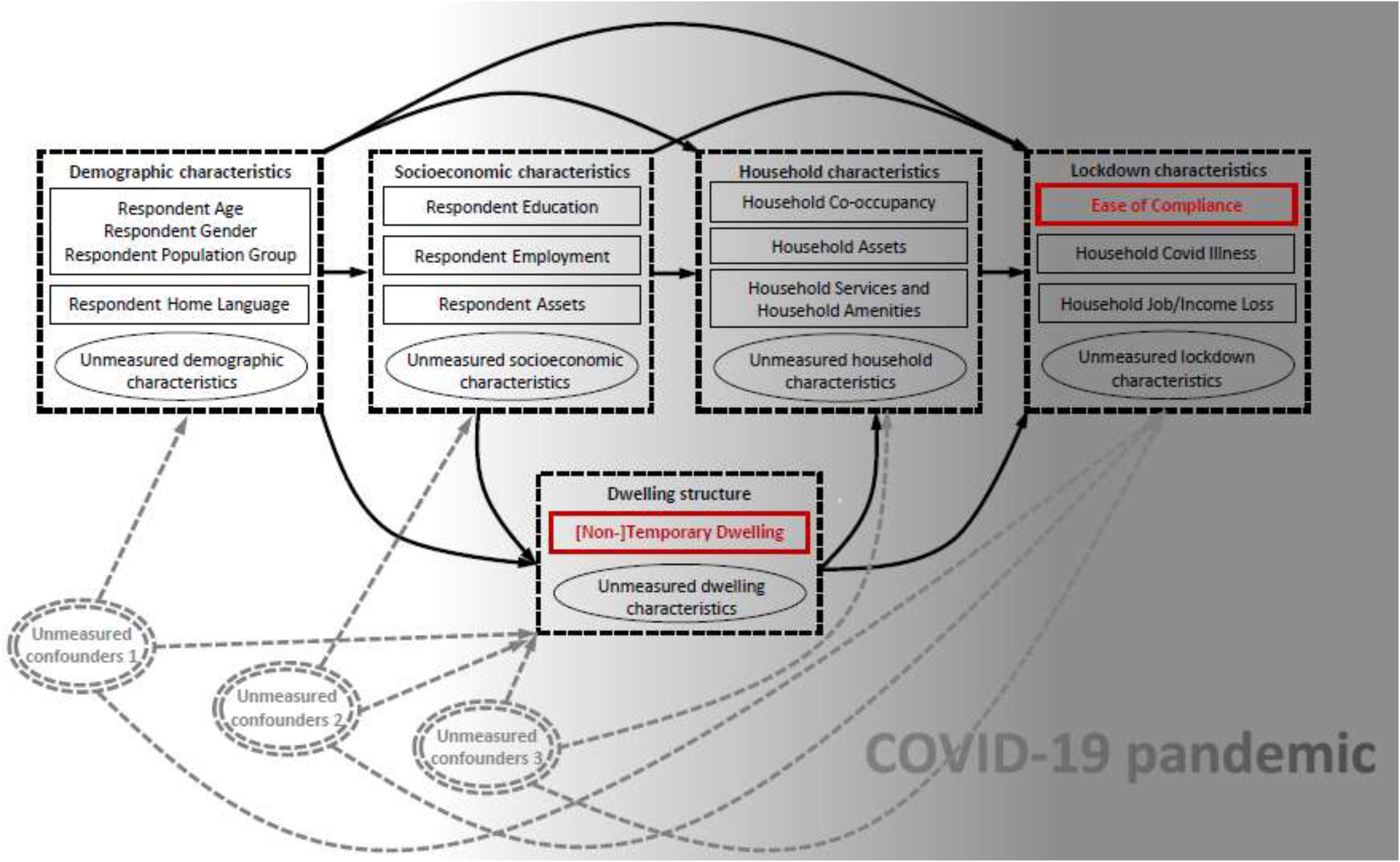
A theoretical causal path diagram, drawn in the form of a directed acyclic graph (DAG) in which each of the measured (rectangles) and unmeasured (ellipses) sets of variables of relevance to the present study have been arranged in their hypothesised temporal sequence (from left to right), with each preceding variable assumed to act as a probabilisitic cause of all subsequent variables. The two specified outcomes (‘[Non-]Temporary Dwelling’ and ‘Ease of Compliance’) have been indicated in red. Three examples of the many different unmeasured sets of covariates (indicated by ellipses with double outlines) likely to contribute confounder bias to estimates of the focal relationships examined in the present study, have been included to emphasise their potential impact.

While this approach to causal inference using observational (i.e. non-experimental) data helps to reduce the impact of bias from *measured* confounders (by ensuring analyses are conditioned thereon), the estimates generated are still likely to be biased,^43^ not least as a result of: residual confounding (associated with non-random measurement error and imprecision in the ascertainment of the variables concerned); unacknowledged/unadjusted confounding (caused by a failure to adjust for unknown/unmeasured/latent confounders); and collider bias (resulting from: endogenous selection bias;^44-46^ or inappropriate conditioning on mediators and/or consequences of the outcome – often as a result of their misclassification as potential/candidate confounders^42^).

In an effort to reduce residual confounding, we carefully examined the responses provided to each of the items available for consideration as potential/candidate confounders to eliminate any sampling/measurement-related error generated by respondents with missing data values, and by overlapping/indiscrete answer options/categories – the first through case-wise deletion of respondents with missing data; and the second through re-categorisation (although both of these steps would have nonetheless introduced alternative sources of bias and imprecision; particularly through selection bias and a loss of information, respectively).

There was less scope to address unadjusted confounding in the design of our analyses, not least given the finite number of items in the AB-R8 survey instrument and its emphasis on self-reported items (and the predominance of opinion-based items) that can be challenging, if not impossible, to interpret as phenomenological events (or crystallising processes) amenable to temporal positioning with respect to any given exposure-outcome dyad. Nonetheless, to acknowledge the potential role that such characteristics might play as unacknowledged or unmeasured confounders, we included three of the innumerable possible sets of unmeasured confounders within our theoretical causal path diagram (Figure 1) to emphasise the (unadjusted confounding) bias they might impose on any estimates of the causal relationships examined in the present study.

Finally, since some of the potential/candidate confounders selected for adjustment comprised features of respondents or households that were subject to change over time (i.e. ‘time-variant’ variables), we sought to address uncertainty regarding precisely when these characteristics might have crystallised (as/when measured by the AB-R8 survey) by conducting sensitivity analyses with confounder/covariate adjustment sets containing potential/candidate confounders considered more vs. less likely to have themselves been affected by COVID-19. The former included: respondent employment status; and respondent/household assets, both of which might plausibly provide indicators of pre-, intra- or post-pandemic socioeconomic vulnerability/mobility.

Notwithstanding our efforts to address these three potential sources of bias when estimating causal effects from observational/non-experimental data, it is important to stress that these efforts are very unlikely to have been completely successful.^42,44^ Similarly, because incompletely representative/non-probabilistic sampling – which is common to most surveys involving relatively small samples of voluntary/consenting participants (such as the AB-R8 survey) – can invoke endogenous selection bias (another form of ‘collider bias’),^45,46^ even carefully theorised causal path diagrams and the careful selection, measurement/parameterisation and statistical adjustment of potential/candidate confounders may not eliminate the risk of generating biased causal estimates from analyses of observational data. For these reasons, the findings generated by the present study remain speculative and warrant careful examination, replication and further exploration.

### Selection of exposure, outcome and ‘candidate’ confounder variables

The two principal outcomes of interest examined in the present study were derived from: an item situated in the final section of the original AB-R8 questionnaire containing fieldworker-generated observations and assessments; and an item included in the supplementary (COVID-19) module attached to the AB-R8 questionnaire in those countries where data collection had been suspended or postponed as a result of the COVID-19 pandemic, and was only resumed following the first (or subsequent) waves of infection (and associated lockdown restrictions). South Africa was one such country, where the AB-R8 survey was conducted between 2 May and 10 June 2021, when the country was in the process of moving from “Alert Level 1” (which at that time included: the closure of 33/53 land border crossings; an overnight curfew; the closure of nightclubs and specified opening/closing hours for public venues; limits on the size of public and private gatherings/events [and a ban on spectators at sporting events]; limits on the distances travelled for work or private purposes; working from home wherever possible; a ban on alcohol consumption in public places; social distancing; the provision of hand sanitisers in public venues; and the wearing of face masks in any ‘public place’)^47,48^ to “Alert Level 2” (in which, *inter alia*, the limits on the size of public and private gatherings/events were further reduced).^49^

The first of these two AB-RA items required the fieldworker to answer the question: *“In what type of shelter does the respondent live?”* with seven pre-categorised answers, namely: *“Non-traditional/Formal house”, “Traditional house/Hut”*; *“Temporary structure/shack”*; *“Flat in a block of flats”*; *“Single room in a larger dwelling structure or backyard”*; *“Hostel in an industrial compound or farming compound”*; or *“Other”*. The second item involved asking respondents (i.e. household key informants): *“How easy or difficult was it for you and your household to comply with the lockdown or curfew restrictions imposed by the government?”* for which there were four explicit answers available (*“Very easy”*; *“Easy”*; *“Difficult”*; or *“Very difficult*”) and three implicit options used to code unprompted and less definitive answers (*“Neither easy nor difficult”*; *“I/we did not comply”*; and *“Don’t know”*). The distribution of responses to each of these items was carefully examined to generate binary categorical variables that sought to balance the distribution of responses with the conceptual integrity of the answers that each provided (as described under ‘Data preparation and statistical analysis’, below; see also Section 1 of the Supplementary Materials).

The variables examined as putative determinants, correlates and consequences of these two outcomes – and as potential/candidate confounders when subsequent variables were specified as the exposure – were selected from amongst those items included in the AB-R8 survey instrument that focussed primarily on phenomenological characteristics (i.e. sociodemographic, economic, household and COVID-19 related features that were least likely to be vulnerable to reporting bias or to have changed substantively [i.e. ‘re-crystallised’] as a result of illness or job/business/income loss during the pandemic). While this meant that a large proportion of the (more opinion-based) items included in the AB-R8 survey instrument had to be discounted as suitable for use as exposures (or potential/candidate confounders), there were a sizeable number of more phenomenological items (ten in all) considered relevant to: the sociodemographic characteristics of respondents (age, gender, and ‘Population Group’ classification) and households (the primary language spoken in the home); and the socioeconomic position of both respondents (educational attainment) and households (type of dwelling, number of adult residents and household utilities, services and amenities), that were considered unlikely to have changed in the 14-15 months from the onset of the COVID-19 pandemic (in March 2020) to when South Africa’s AB-R8 survey took place (in May-June 2021). While responses to items on these characteristics were therefore considered ‘time-invariant’ (and to have occurred at discrete points before each of the present study’s specified outcomes), there were a number of additional, *ostensibly* phenomenological, criteria (including: respondent employment; 12 measures of discrete respondent/household assets; and 2 measures of COVID-19 related impacts on illness and job/business/income loss) that were likely to have been more susceptible to change during/following the onset of the pandemic, and were therefore considered ‘time-variant’ (and to have potentially crystallised – as/when measured – *after both* of the principal outcomes examined in the present study). To address the risk that these 14 variables might not constitute genuine confounders (but instead might act as colliders, whether as: consequences of the outcome; or mediators between each of the exposure-outcome dyads examined), we undertook the sensitivity analyses described earlier (see Table S2.1 and S2.2 in Section 2 of the Supplementary Materials) using confounder/covariate adjustment sets that excluded: respondent employment status; and respondent/household assets – viewing these instead as likely indicators of intra/post-pandemic fluidity in socioeconomic position (as opposed to definitive measures of pre-pandemic employment/wealth). These analyses are examined in greater detail below (see: Results - Multivariable statistical analyses - Determinants, correlates and consequences of ease of compliance with lockdown restrictions).

### Data preparation and statistical analyses

In preparation for our analyses, the distribution of responses to all 24 of the items selected as putative determinants, correlates and/or consequences of each of the two specified outcomes – or as potential/candidate confounders in any of the exposure-outcome dyads involved – was carefully examined to facilitate their re-categorisation into coherent analytical variables (see Section 1 of the Supplementary Materials). This included reducing each of the specified outcomes to binary variables for analysis using logistic regression analysis, in which the categories selected were determined at, or as close as possible to, the median value.

Standard descriptive statistics (frequencies with percentages) were used to summarise the responses obtained for each of the 26 variables (i.e. 24 covariates and 2 specified outcomes) examined in the present study. Respondents who were ineligible/unable/unwilling to answer (or did not know the answer to) any of the survey items required to generate these data were excluded from the (sub)sample of respondents subsequently included in the ‘complete case analyses’ that followed. These analyses involved univariable and multivariable logistic regression models designed with reference to the theoretical causal path diagram summarised in Figure 1, in which the postulated temporal sequence of, and probabilistic causal relationships between, each of these variables was used to select covariates likely to have acted as potential confounders for each of the exposure-outcome dyads examined. The results of these (unadjusted and confounder adjusted) models are presented as odds ratios (OR) with 95% confidence intervals in parentheses (95%CI).

## Results

### Sample characteristics

The re-categorised variables derived from each of the 26 AB-R8 items have been summarised in Table 1 (below). This indicates that most respondents (1,381; 86.3%) provided complete data on all 26 variables, while a modest number (219; 13.7%) provided responses to one or more of the survey items (e.g. “Don’t know”, “Refused”, or “Not applicable”) that resulted in missing data values. Given the risk of endogenous selection bias in analyses that seek causal inference from unrepresentative samples^45^ – a risk that can already be high in studies dependent on fallible and incompletely probabilistic sampling techniques (such as household surveys conducted under exigent circumstances)^49-51^ – the distribution of responses obtained from participants providing complete data on all 26 variables was compared to those of participants who had not (Table 1). This comparison provided some reassurance that the former (the ‘complete case [sub]sample’) displayed sociodemographic and economic characteristics (at both the individual- and household-level) that were broadly comparable to the latter. In particular, it was reassuring that the proportion of respondents in each (sub)sample who were resident in non-temporary structures (i.e. houses/flats, rooms or hostels) was very similar (91.0% vs. 87.8%); as was the proportion of those who had found it “Difficult” (or worse) to comply with lockdown restrictions (34.7% vs. 40.3%); and there was also little difference in the proportion of households who had experienced COVID-19 related illness (19.8% vs. 17.5%) or job/business/income loss (34.1% vs. 32.3%; see Table 1).

**Table 1.** The distribution of the 26 re-categorised items from the AB-R8 survey amongst: (i) the 1,381 (86.3%) South African respondents who provided answers to all 26 items; and (ii) the 219 respondents (13.7 %) with missing data on one or more of these variables.

| Variable |  | Respondents with complete data<br>(n=1,381) |  | Respondents with incomplete data<br>(n≤219) |  |
| --- | --- | --- | --- | --- | --- |
|  |  | n | (%) | n | (%) |
| **Respondent Age** | 18-25yr (3) | 325 | 23.5 | 41 | 18.8 |
|  | 26-35yr (2) | 377 | 27.3 | 68 | 31.2 |
|  | 36-50yr (1) | 390 | 28.2 | 63 | 28.9 |
|  | 51-90yr (0) | 289 | 20.9 | 46 | 21.1 |
| **Respondent Gender** | Female (0) | 697 | 50.5 | 104 | 47.5 |
|  | Male (1) | 684 | 49.5 | 115 | 52.5 |
| **Respondent 'Population Group' classification** | 'Black/African' (0) | 971 | 70.3 | 133 | 60.7 |
|  | 'White/European' (1) | 139 | 10.1 | 29 | 13.2 |
|  | 'Coloured/Mixed race' (2) | 206 | 14.9 | 38 | 17.4 |
|  | 'South/East Asian' (3) | 65 | 4.7 | 19 | 8.7 |
| **Respondent Home Language** | Non-European (1) | 1,051 | 76.1 | 164 | 74.9 |
|  | European (0) | 330 | 23.9 | 55 | 25.1 |
| **Respondent Education** | Less than complete primary (4) | 140 | 10.1 | 24 | 11.6 |
|  | Complete primary but less than complete secondary (1) | 397 | 28.8 | 55 | 26.6 |
|  | Secondary complete (0) | 489 | 35.4 | 56 | 27.1 |
|  | Non-university post-secondary (3) | 151 | 10.9 | 24 | 11.6 |
|  | Some university or more (2) | 204 | 14.8 | 48 | 23.2 |
| **Respondent Employment** | No (looking) (0) | 534 | 38.7 | 74 | 36.8 |
|  | No (not looking) (2) | 334 | 24.2 | 47 | 23.4 |
|  | Yes, part time (3) | 173 | 12.5 | 24 | 11.9 |
|  | Yes, full time (1) | 340 | 24.6 | 56 | 27.9 |
| **Respondent Assets** | Radio – Yes (1) | 943 | 68.3 | 159 | 72.6 |
|  | TV – Yes (1) | 955 | 69.2 | 165 | 75.3 |
|  | Motor Vehicle – Yes (1) | 381 | 27.6 | 72 | 32.9 |
|  | Computer – Yes (1) | 480 | 34.8 | 102 | 46.5 |
|  | Account – Yes (1) | 1,121 | 81.2 | 184 | 84.0 |
|  | Mobile – Yes (1) | 1,263 | 91.5 | 189 | 86.3 |
| **Residence in [Non-]Temporary Structure** | Formal house/flat (0) | 1,208 | 87.5 | 175 | 85.4 |
|  | Temporary structure/shack (1) | 125 | 9.1 | 25 | 12.2 |
|  | Single room/hostel (2) | 48 | 3.5 | 5 | 2.4 |
| **Household Co-occupancy** | One adult in household (0) | 599 | 43.4 | 81 | 37.0 |
|  | Two adults in household (1) | 354 | 25.6 | 50 | 22.8 |
|  | More than two adults in household (2) | 428 | 31.0 | 88 | 40.2 |
| **Household asset ownership** | Radio – Yes (1) | 1,174 | 85.0 | 186 | 84.9 |
|  | TV – Yes (1) | 1,270 | 92.0 | 194 | 88.6 |
|  | Motor Vehicle – Yes (1) | 652 | 47.2 | 111 | 50.7 |
|  | Computer – Yes (1) | 654 | 47.4 | 120 | 54.8 |
|  | Account – Yes (1) | 1,244 | 90.1 | 198 | 90.4 |
|  | Mobile – Yes (1) | 1,326 | 96.0 | 203 | 92.7 |
| **Household Electricity Supply** | Connected to grid (0) | 1,257 | 91.0 | 189 | 89.6 |
|  | Not connected to grid (1) | 124 | 9.0 | 22 | 10.4 |
| **Household Water Supply** | Water inside dwelling (0) | 733 | 53.1 | 123 | 63.7 |
|  | Water inside compound (1) | 395 | 28.6 | 35 | 18.1 |
|  | Water outside compound (2) | 253 | 18.3 | 35 | 18.1 |
| **Household Toilet Access** | Private inside dwelling (0) | 694 | 50.3 | 116 | 53.7 |
|  | Private inside compound (1) | 407 | 29.5 | 46 | 21.3 |
|  | Outside compound (shared/none available) (2) | 280 | 20.3 | 54 | 25.0 |
| **Household Ease of Lockdown Compliance** | Less than “Difficult” (0) | 497 | 34.7 | 27 | 40.3 |
|  | “Difficult” or worse (1) | 902 | 65.3 | 40 | 59.7 |
| **Household COVID-19 related Illness** | Did not become ill (0) | 1,108 | 80.2 | 52 | 82.5 |
|  | Became ill (1) | 273 | 19.8 | 11 | 17.5 |
| **Household COVID-19 related Job/Business/Income loss** | No loss (0) | 910 | 65.9 | 44 | 67.7 |
|  | Lost job/business/income (1) | 471 | 34.1 | 21 | 32.3 |

However, there were nonetheless some more substantive differences evident in the sociodemographic distribution of the complete case (sub)sample, with 10% more respondents classified as ‘Black/African’ (70.3% vs. 60.7%) and only half the proportion classified as ‘South/East Asian’ (4.7% vs. 8.7%) when compared to those with missing data. Given South Africa’s enduring legacy of structural and socioeconomic inequality along quasi-racialised ‘Population Group’ lines,^21^ these differences might explain the lower educational attainment of respondents in the complete case (sub)sample (e.g. 14.8% vs. 23.2% having completed at least some university/tertiary education) and the lower proportion of these respondents who owned all but one of the six personal assets (the exception being a mobile phone). Despite these trends, multiple co-occupancy was actually lower amongst households included in the complete case (sub)sample (with only 31.0% vs. 40.2% of these households occupied by more than two adults); and there was little evidence of any substantive differences in household asset ownership or in access to household services (the notable exception being the proportion of households with piped water inside their dwelling: 53.1% vs. 63.7%; see Table 1).

Notwithstanding these differences (and the potential risk of endogenous selection/collider bias they might pose),^45^ the analyses that follow rely solely on those 1,381 respondents for whom data were available on all 26 of the variables examined, and the results of these analyses therefore need to be interpreted with a degree of caution from a causal inference perspective.^44,45,49^ This (sub)sample of adult respondents comprised a similar number of men and women, with a median age of 35 (range: 18-90), most of whom were classified as ‘Black/African’ (70.3%), and with far fewer classified as ‘Coloured/mixed race’ (14.9%), ‘White/European’ (10.1%) or ‘South/East Asian’ (4.7%). Most (76.1%) spoke non-European languages at home (the majority of which were indigenous South African languages; see Section 1 of the Supplementary Materials); and although 61.1% had completed secondary education (or above), only around a third (37.1%) reported they had current employment that paid a cash income, and around a third of these (12.5%) were only employed part-time. As such, the complete case (sub)sample of South African respondents included in the analyses that follow are characterised by high levels of under-employment, and this is likely to have a substantial bearing on the proportion who reported that they, or someone in their household, had (temporarily or permanently) lost their income/job/business as a result of COVID-19 (34.6%) – particularly if, as seems likely, a substantial proportion of those who reported that they were under-employed at the time the AB-R8 survey took place had lost employment/income *as a result of* COVID-19. Under such circumstances (and as described earlier), this employment variable (and its associated impact on the assets owned by respondents and their households) seems very likely to constitute a potential *consequence* of COVID-19 rather than always being a preceding *determinant* of either residence in a temporary structure/shack or the ease of lockdown compliance.

### Multivariable statistical analyses

To address the possibility that individual- and household-level socioeconomic characteristics (i.e. respondent employment and individual/household assets) might constitute consequences (as opposed to determinants) of residence in a temporary structure/shack and/or ease of lockdown compliance – and might therefore act as mediators/colliders rather than genuine confounders in the sociodemographic patterning of either specified outcome, additional sensitivity analyses were undertaken in which employment and personal/household assets were removed from the covariate adjustment sets used to mitigate the effect of confounder bias (see Table S2.1 and S2.2 in Section 2 of the Supplementary Materials). These additional analyses mirrored the statistical models used to estimate the total causal effects of each of the remaining 25 (sociodemographic, economic, household and COVID-19 related) variables selected for examination as potential determinants, correlates or consequences of residence in a temporary structure/shack and/or ease of lockdown compliance (see Table 2 and 3). The first of these sets of models (summarised in the first column of Table 2 and 3) adjusted for none of the preceding (candidate) covariates considered potential confounders; while the second set (summarised in the second column of each Table) adjusted only for those individual- and household-level sociodemographic and economic covariates considered likely to have occurred (or crystallised) *before* the specified outcome.

**Table 2.** The sociodemographic and economic determinants of residence in a temporary structure/shack vs. a non-temporary/permanent structure (i.e. a house/flat, room or hostel); and the relationship between residence in a (non-)temporary structure/shack and a number of: household-level characteristics (including co-occupancy and household assets, services and amenities); and COVID-19 related phenomena (ease of lockdown compliance, illness and job/business income loss) – both before (Column 2.1) and after (Column 2.2) adjustment for any preceding potential/candidate confounders. All results are presented as odds ratios (ORs) with 95% confidence intervals in parentheses (95%CI).

|  |  |  |  |
| --- | --- | --- | --- |
| **Specified outcome:** | Residence in a temporary structure/shack vs. a non-temporary/permanent structure (house/flat, room or hostel) – dichotomous |  |  |
| Outcome referent (0): | Non-temporary structure (house/flat, room or hostel) |  |  |
| Outcome contrast (1): | Temporary structure/shack |  |  |
| **Specified exposures:** |  | **Covariate adjustment set** |  |
|  |  | **Column 2.1<sup>1</sup>** | **Column 2.2<sup>2</sup>** |
|  |  | None | Any preceding covariates |
|  |  | OR (95%CI) | OR (95%CI) |
| **Covariates considered likely to precede (and be potential determinants of) residence in a (non-)temporary structure:<sup>3</sup>** |  |  |  |
| **Respondent Age** | 18-25yr (3) | **3.25 (1.62,6.49)** | **3.23 (1.60,6.50)** |
|  | 26-35yr (2) | **3.17 (1.60,6.27)** | **3.05 (1.54,6.06)** |
|  | 36-50yr (1) | **2.49 (1.24,4.99)** | **2.76 (1.37,5.55)** |
|  | 51-90yr (0) | Referent | Referent |
| **Respondent Gender** | Female (0) | Referent | Referent |
|  | Male (1) | 1.11 (0.77,1.61) | 1.11 (0.77,1.61) |
| **Respondent ‘Population Group’ classification** | ‘Black/African’ (0) | Referent | Referent |
|  | ‘White/European’ (1) | **0.12 (0.03,0.49)** | **0.11 (0.03,0.46)** |
|  | ‘Coloured/Mixed race’ (2) | 0.79 (0.47,1.33) | 0.78 (0.46,1.35) |
|  | ‘South/East Asian’ (3) | 1 (Empty) | 1 (Empty) |
| **Respondent Home Language** | Non-European (0) | Referent | Referent |
|  | European (1) | 0.66 (0.41,1.06) | 1.09 (0.65,1.82) |
| **Respondent Education** | Less than complete primary (4) | 1.19 (0.65,2.16) | **2.01 (1.04,3.88)** |
|  | Less than complete secondary (1) | 1.35 (0.89,2.06) | 1.53 (0.99,2.35) |
|  | Secondary complete (0) | Referent | Referent |
|  | Non-university post-secondary (3) | **0.38 (0.16,0.91)** | **0.38 (0.16,0.91)** |
|  | Some university or more (2) | **0.18 (0.07,0.52)** | **0.24 (0.08,0.67)** |
| **Respondent Employment** | No job (looking) (0) | Referent | Referent |
|  | No (not looking) (2) | **0.47 (0.29,0.76)** | 0.66 (0.39,1.12) |
|  | Yes (part time) (3) | **0.47 (0.25,0.89)** | 0.57 (0.29,1.09) |
|  | Yes (full time) (1) | **0.33 (0.19,0.57)** | **0.54 (0.30,0.96)** |
| **Respondent Radio** | Do not personally own (0) | Referent | Referent |
|  | Personally own (1) | **0.65 (0.44,0.94)** | 0.77 (0.51,1.16) |
| **Respondent Television** | Do not personally own (0) | Referent | Referent |
|  | Personally own (1) | **0.45 (0.31,0.65)** | **0.57 (0.38,0.85)** |
| **Respondent Motor Vehicle** | Do not personally own (0) | Referent | Referent |
|  | Personally own (1) | **0.26 (0.14,0.47)** | **0.48 (0.25,0.92)** |
| **Respondent Computer** | Do not personally own (0) | Referent | Referent |
|  | Personally own (1) | **0.35 (0.22,0.57)** | **0.55 (0.32,0.93)** |
| **Respondent Bank Account** | Do not personally own (0) | Referent | Referent |
|  | Personally own (1) | 0.87 (0.55,1.37) | 1.25 (0.76,2.05) |
| **Respondent Mobile Phone** | Do not personally own (0) | Referent | Referent |
|  | Personally own (1) | **0.41 (0.25,0.69)** | **0.43 (0.25,0.76)** |
| **Covariates considered likely to be coterminous with (or determined by) residence in a [non-]temporary structure:** |  |  |  |
| **Household Co-occupancy** | One adult in household (0) | Referent | Referent |
|  | Two adults in household (1) | 1.51 (0.95,2.39) | **1.77 (1.09,2.88)** |
|  | More than two adults in household (2) | 1.52 (0.98,2.35) | **1.87 (1.17,3.00)** |
| **Household Radio** | No household member owns (0)<br>Household member owns (1) | Referent<br>**0.43 (0.28,0.65)** | Referent<br>**0.32 (0.16,0.62)** |
| **Household Television** | No household member owns (0)<br>Household member owns (1) | Referent<br>**0.21 (0.13,0.33)** | Referent<br>**0.23 (0.12,0.43)** |
| **Household Motor Vehicle** | No household member owns (0)<br>Household member owns (1) | Referent<br>**0.24 (0.15,0.38)** | Referent<br>**0.31 (0.16,0.58)** |
| **Household Computer** | No household member owns (0)<br>Household member owns (1) | Referent<br>**0.38 (0.25,0.58)** | Referent<br>0.62 (0.33,1.18) |
| **Household Bank Account** | No household member owns (0)<br>Household member owns (1) | Referent<br>0.67 (0.39,1.16) | Referent<br>0.53 (0.22,1.31) |
| **Household Mobile Phone** | No household member owns (0)<br>Household member owns (1) | Referent<br>**0.30 (0.16,0.57)** | Referent<br>0.42 (0.15,1.19) |
| **Household Electricity Supply** | Connected to grid (0)<br>Not connected to grid (1) | Referent<br>**12.89 (8.40,19.77)** | Referent<br>**11.80 (7.03,19.81)** |
| **Household Water Supply** | Water inside dwelling (0)<br>Water inside compound (1)<br>Water outside compound (2) | Referent<br>**3.12 (1.92,5.06)**<br>**6.13 (3.79,9.92)** | Referent<br>**2.82 (1.67,4.77)**<br>**3.58 (2.09,6.13)** |
| **Household Toilet Access** | Private inside dwelling (0)<br>Private inside compound (1)<br>Outside compound (shared/none available) (2) | Referent<br>**6.48 (3.66,11.49)**<br>**10.36 (5.82,18.44)** | Referent<br>**4.19 (2.22,7.90)**<br>**5.78 (3.01,11.11)** |
| **Household Ease of Lockdown Compliance** | Less than “Difficult” (0)<br>“Difficult” or worse (1) | Referent<br>**1.61 (1.06,2.45)** | Referent<br>0.99 (0.60,1.63) |
| **Household COVID-19 related Illness** | Did not become ill (0)<br>Became ill (1) | Referent<br>0.61 (0.36,1.04) | Referent<br>1.42 (0.75,2.67) |
| **Household COVID-19 related Job/Business/Income loss** | No loss (0)<br>Lost job/business/income (1) | Referent<br>1.10 (0.75,1.61) | Referent<br>1.34 (0.83,2.17) |

<sup>1</sup>Column 2.1: No adjustment for potential/preceding candidate confounding covariates.
<sup>2</sup>Column 2.2: Adjustment only for any preceding candidate/potential confounding covariates (and not for coterminous covariates in square parentheses – [...]; with the exception of Respondent Age, Gender, ‘Population Group’ and Home Language), as listed in the following sequence: Respondent Age, Gender, ‘Population Group’, Home Language, Respondent Education; Respondent Employment; [Respondent Assets – Radio, Television, Motor Vehicle, Computer, Bank Account, Mobile Phone]; Household Occupancy; [Household Assets – Radio, Television, Motor Vehicle, Computer, Bank Account, Mobile Phone]; [Household Services and Amenities – Electricity, Water, Toilet]; [Lockdown Characteristics – Ease of Compliance, Household COVID-19 Illness, Job/Business/Income Loss].
<sup>3</sup>Alternating white/grey shading indicates groups of covariates considered coterminous (i.e. occurring or crystallising at around the same time, given the questions/items used to ascertain these within the AB-R8 survey).

**Table 3.** The sociodemographic and economic determinants of ease of compliance with COVID-19 restrictions; and the relationship between ease of compliance and COVID-19 related illness and job/business/income loss; both before (Column 3.1) and after (Column 3.2) adjustment for any preceding potential/candidate confounders. All results are presented as odds ratios (ORs) with 95% confidence intervals in parentheses (95%CI).

| Specified outcome: |  | Self-reported ease of compliance with lockdown restrictions – dichotomous |  |
| --- | --- | --- | --- |
| Outcome referent (0): |  | Less than “Difficult” |  |
| Outcome contrast (1): |  | “Difficult” or worse |  |
| Specified exposures: |  | Covariate adjustment set |  |
|  |  | Column 3.1<sup>1</sup> | Column 3.2<sup>2</sup> |
|  |  | None | Any preceding covariates |
|  |  | OR (95%CI) | OR (95%CI) |
| Covariates considered likely to precede (and be potential determinants of) ease of compliance with lockdown:<sup>3</sup> |  |  |  |
| Respondent Age | 18-25yr (3) | 1.35 (0.97,1.87) | 1.35 (0.97,1.89) |
|  | 26-35yr (2) | **1.57 (1.14,2.16)** | **1.55 (1.12,2.15)** |
|  | 36-50yr (1) | 1.23 (0.90,1.68) | 1.32 (0.96,1.81) |
|  | 51-90yr (0) | Referent | Referent |
| Respondent Gender | Female (0) | Referent | Referent |
|  | Male (1) | 1.11 (0.89,1.39) | 1.13 (0.90,1.41) |
| Respondent ‘Population Group’ classification | ‘Black/African’ (0) | Referent | Referent |
|  | ‘White/European’ (1) | **0.46 (0.32,0.66)** | **0.45 (0.30,0.67)** |
|  | ‘Coloured/Mixed race’ (2) | 0.78 (0.57,1.07) | 0.78 (0.57,1.08) |
|  | ‘South/East Asian’ (3) | **0.53 (0.32, 0.88)** | **0.55 (0.30, 0.98)** |
| Respondent Home Language | Non-European (0) | Referent | Referent |
|  | European (1) | **0.76 (0.59,0.99)** | 1.02 (0.74,1.40) |
| Respondent Education | Less than complete primary (4) | 0.98 (0.66,1.47) | 1.17 (0.76,1.81) |
|  | Less than complete secondary (1) | 1.14 (0.86,1.52) | 1.20 (0.89,1.61) |
|  | Secondary complete (0) | Referent | Referent |
|  | Non-university post-secondary (3) | 0.71 (0.49,1.04) | 0.72 (0.49,1.05) |
|  | Some university or more (2) | **0.55 (0.40,0.77)** | **0.62 (0.44,0.87)** |
| Respondent Employment | No job (looking) (0) | Referent | Referent |
|  | No (not looking) (2) | **0.63 (0.47,0.85)** | 0.75 (0.55,1.03) |
|  | Yes (part time) (3) | **0.62 (0.43,0.89)** | 0.69 (0.48,1.01) |
|  | Yes (full time) (1) | **0.55 (0.42,0.74)** | **0.72 (0.52,0.99)** |
| Respondent Radio | Do not personally own (0) | Referent | Referent |
|  | Personally own (1) | 0.85 (0.67,1.08) | 0.94 (0.73,1.22) |
| Respondent Television | Do not personally own (0) | Referent | Referent |
|  | Personally own (1) | 0.92 (0.72,1.17) | 1.10 (0.84,1.42) |
| Respondent Motor Vehicle | Do not personally own (0) | Referent | Referent |
|  | Personally own (1) | **0.56 (0.44,0.72)** | 0.76 (0.57,1.02) |
| Respondent Computer | Do not personally own (0) | Referent | Referent |
|  | Personally own (1) | **0.59 (0.47,0.74)** | **0.71 (0.54,0.93)** |
| Respondent Bank Account | Do not personally own (0) | Referent | Referent |
|  | Personally own (1) | 0.86 (0.64,1.15) | 0.99 (0.72,1.37) |
| Respondent Mobile Phone | Do not personally own (0) | Referent | Referent |
|  | Personally own (1) | 1.32 (0.90,1.95) | 1.46 (0.98,2.19) |
| Residence in a Temporary/Non-temporary Structure | House/flat (0) | Referent | Referent |
|  | Temporary structure/shack (1) | **1.61 (1.06,2.45)** | 1.20 (0.78,1.87) |
|  | Single room/hostel (2) | 1.11 (0.60,2.04) | 0.88 (0.47,1.66) |
| Household Co-occupancy | One adult in household (0) | Referent | Referent |
|  | Two adults in household (1) | 0.88 (0.66,1.15) | 0.92 (0.69,1.22) |
|  | More than two adults in household (2) | 0.86 (0.67,1.12) | 0.90 (0.67,1.18) |
| **Household Radio** | No household member owns (0)<br>Household member owns (1) | Referent<br>0.84 (0.61,1.15) | Referent<br>1.00 (0.65,1.54) |
| **Household Television** | No household member owns (0)<br>Household member owns (1) | Referent<br>1.16 (0.78,1.73) | Referent<br>1.38 (0.85,2.24) |
| **Household Motor Vehicle** | No household member owns (0)<br>Household member owns (1) | Referent<br>**0.60 (0.48,0.75)** | Referent<br>0.81 (0.60,1.11) |
| **Household Computer** | No household member owns (0)<br>Household member owns (1) | Referent<br>**0.60 (0.48,0.76)** | Referent<br>0.86 (0.60,1.24) |
| **Household Bank Account** | No household member owns (0)<br>Household member owns (1) | Referent<br>1.13 (0.78,1.63) | Referent<br>1.55 (0.89,2.69) |
| **Household Mobile Phone** | No household member owns (0)<br>Household member owns (1) | Referent<br>1.08 (0.62,1.89) | Referent<br>0.77 (0.36,1.65) |
| **Household Electricity Supply** | Connected to grid (0)<br>Not connected to grid (1) | Referent<br>1.39 (0.92,2.09) | Referent<br>1.29 (0.81,2.03) |
| **Household Water Supply** | Water inside dwelling (0)<br>Water inside compound (1)<br>Water outside compound (2) | Referent<br>**1.30 (1.00,1.68)**<br>**1.63 (1.19,2.22)** | Referent<br>1.08 (0.82,1.43)<br>1.28 (0.90,1.81) |
| **Household Toilet Access** | Private inside dwelling (0)<br>Private inside compound (1)<br>Outside compound (shared/none available) (2) | Referent<br>**1.37 (1.06,1.77)**<br>**2.18 (1.59,2.99)** | Referent<br>1.02 (0.76,1.38)<br>**1.55 (1.08,2.24)** |
| **Covariates considered likely to be coterminous with (or consequences of) ease of lockdown compliance:<sup>2</sup>** |  |  |  |
| **Household COVID-19 related Illness** | Did not become ill (0)<br>Became ill (1) | Referent<br>0.83 (0.63,1.09) | Referent<br>1.05 (0.79,1.41) |
| **Household COVID-19 related Job/Business/Income loss** | No loss (0)<br>Lost job/business/income (1) | Referent<br>**1.88 (1.47,2.40)** | Referent<br>**1.87 (1.44,2.43)** |

<sup>1</sup>Column 3.1: No adjustment for potential/preceding candidate confounding covariates.
<sup>2</sup>Column 3.2: Adjustment only for any preceding candidate/potential confounding covariates (and not for coterminous covariates in square parentheses – [...]; with the exception of Respondent Age, Gender, ‘Population Group’ and Home Language), as listed in the following sequence: Respondent Age, Gender, ‘Population Group’, Home Language, Respondent Education; Respondent Employment; [Respondent Assets – Radio, Television, Motor Vehicle, Computer, Bank Account, Mobile Phone]; [Residence in a (Non-)Temporary Structure, Household Occupancy]; [Household Assets – Radio, Television, Motor Vehicle, Computer, Bank Account, Mobile Phone]; [Household Services and Amenities – Electricity, Water, Toilet]; [Lockdown Characteristics – Household COVID-19 Illness, Job/Business/Income Loss].
<sup>3</sup>Alternating white/grey shading indicates groups of covariates considered coterminous (i.e. occurring or crystallising at around the same time, given the questions/items used to ascertain these within the AB-R8 survey).

#### Determinants, correlates and consequences of residence in a (non-)temporary structure/shack

Table 2 summarises both: the sociodemographic and economic determinants of residence in a temporary structure/shack (as compared to a non-temporary/permanent structure, such as a house/flat, room or hostel); and the relationships evident between residence in a temporary structure/shack and a number of household-level characteristics (including co-occupancy and household assets, services and amenities) and COVID-19 related phenomena (ease of lockdown compliance, illness and job/business income loss). These analyses confirm that residence in a temporary structure/shack was far more common amongst respondents aged 50yr or younger (when compared to those aged 51-90yr), but was far less common amongst respondents classified as ‘White/European’ (or indeed ‘South/East Asian’, *none* of whom lived in a temporary structure/shack) than those classified as ‘Black/African’.

Residence in a temporary structure/shack was also far less common amongst respondents who were better educated (and particularly those who had completed some university/tertiary education), and amongst those who were in full or part time employment. Moreover, even those who were unemployed but not looking for work were less likely to live in a temporary structure/shack than those who were looking for work – presumably because the former were able to rely on another source of income (such as a pension, grant, private income or a wealthy/working partner/family member) that was sufficient to cover the cost of living in a non-temporary/formal dwelling (i.e. a house/flat, room or hostel). These socioeconomic patterns were also evident in the much lower odds of residence in a temporary structure/shack amongst those respondents who had the means to own material assets (particularly a television, motor vehicle or computer), even after adjustment for employment status. Meanwhile, *households* that lacked key material assets (particularly a television, motor vehicle or computer) were also far more likely to reside in a temporary structure/shack; as were households that were not connected to the electricity grid, or did not have piped water or private toilet facilities within their own dwelling.

To a large extent, these relationships were only modestly attenuated following adjustment for potential confounders; and excluding respondent employment and respondent/household assets from the confounder adjustment sets in the sensitivity analyses summarised in Table S2.1 (see Section 2 of the Supplementary Materials), indicated that non/adjustment for these (*potentially* time-variant) markers of socioeconomic status had little effect on the relationships observed amongst the putative determinants, correlates and consequences of residence in a temporary structure/shack. Taken together, these analyses indicate that residence in informal structures/shacks is consistently associated with sociodemographic and economic indicators of disadvantage and poverty; and that such dwellings have fewer of the material assets, services and amenities that might otherwise help mitigate the impact of disadvantage and poverty on health, and the vulnerability of their residents to COVID-19 related illness and job/business/income loss (though both these relationships lacked precision before and after adjustment for potential confounders).

#### Determinants, correlates and consequences of ease of compliance with lockdown restrictions

Table 3 summarises both: the sociodemographic and economic determinants of ease of compliance with COVID-19 restrictions; and the relationship between ease of compliance and COVID-19 related illness and job/business/income loss. These analyses reveal that respondents whose households had found it “Difficult” or “Very difficult” to comply with lockdown restrictions (or had been unable to comply with these) were very similar to those who were more likely to reside in temporary structures/shacks (see Table 2). For example, younger respondents were more likely to live in households that found it “Difficult” (or worse) to comply with lockdown restrictions; while those who were classified as ‘White/European’ or ‘South/East Asian’, and those with some university education, who were employed (full or part time), or who owned substantive assets (particularly a motor vehicle or a computer) were far *less* likely to have found it “Difficult” (or harder still) to comply with lockdown restrictions.

For these reasons it may not be surprising that respondents who were residents of temporary structures/shacks were more likely to report that their household had found it “Difficult” (or worse) to comply with lockdown restrictions (OR: 1.61; 95%CI: 1.06,2.45). However, this relationship was substantially attenuated and lost precision following adjustment for preceding (individual-level) sociodemographic and economic confounders (OR: 1,21; 95%CI: 0.78,1.87). Instead, ease of compliance with lockdown was most strongly associated with individual-level sociodemographic/economic characteristics (as summarised above), and with household-level characteristics that were more commonly (though not exclusively) observed amongst residents in a non-temporary/permanent structure (such as: household ownership of a motor vehicle, and/or computer; and piped water/private toilet facilities within the dwelling).

While many of the relationships observed between respondent- and household-level characteristics and ease of compliance with lockdown restrictions were substantively attenuated following adjustment for potential confounders, those for age, ‘Population Group’, education, employment and at least one key personal asset (a computer) retained precision; as did the association with household toilet facilities – where respondents in households lacking a private toilet within their dwelling were far more likely to report that complying with lockdown restrictions had been “Difficult” (or worse), even after adjustment for all 20 preceding variables considered potential confounders (OR: 1.55; 95%CI: 1.08,2.24). Furthermore, the strength and precision of these (confounder adjusted) relationships was largely unaffected when respondent employment and respondent/household assets were excluded from the confounder adjustment sets used in the sensitivity analyses summarised in Table S2.2 (see Section 2 of the Supplementary Materials) – indicating that non/adjustment for these (*potentially* time-variant) markers of socioeconomic status had little effect on the relationships observed amongst the putative determinants, correlates and consequences of ease of compliance with COVID-19 lockdown restrictions.

These relationships therefore reveal that many of the determinants and characteristics of residence in a temporary structure/shack also have a substantive impact on the ease of compliance with South Africa’s COVID-19 lockdown restrictions. As such, the health risks that residents in these households face as a result of their disadvantage and poverty will have been amplified not only by the additional risks, limited assets and inferior services that living in such structures affords, but also by their limited ability to comply with measures intended to reduce the transmission of COVID-19 (i.e. stay at home, maintain personal hygiene and practice social distancing). Indeed, while there was some evidence within the dataset examined in the present study that households who had experienced COVID-19 related illness, or had lost a job, a business or income as a result of the pandemic, were *more* likely to reside in temporary structures/shacks (see Table 2), both of these relationships appeared prone to substantial bias from confounding, and both lacked precision (illness – OR:1.42; 95%CI: 0.75,2.67; job/business/income loss – OR:1.34; 95%CI: 0.83,2.17). Nonetheless, the heightened vulnerability of such households to the ill-effects of COVID-19 and associated lockdown restrictions is evident amongst those that experienced COVID-19 related job/business/income loss who – like respondents who were looking for work – were far more likely to find it “Difficult” (or worse) to comply with lockdown restrictions (even after adjustment for all 23 preceding variables; OR: 1.87; 95%CI: 1.44,2.43; see Table 3).

## Discussion

Our analyses confirm the unequal distribution of sociodemographic and economic circumstances amongst South African households living in temporary structures/shacks.^52,53^ Setting aside the agency and determination evident wherever the poor and underserved have taken the initiative to ‘house themselves’ (as some believe policy makers implicitly concede they must),^6,7,14,23^ the evidence is clear that the residents of these households tend to: come from South Africa’s most disadvantaged ‘Population Group’ (i.e. those classified as ‘Black/African’); have lower educational attainment and fewer personal (and household) assets; be under-employed/looking for work; and have to share water and toilet facilities, while often coping without access to mains electricity.^19,27^ These same factors are also strong determinants, correlates and consequences of compliance with the COVID-19 lockdown restrictions introduced in 2020 (and still in place, though presently at a much lower ‘tier’ than initially implemented). Unsurprisingly, survey respondents living in temporary structures/shacks were much more likely to report that they had not been able to comply with these restrictions (or had found it “Difficult” or “Very difficult” to do so). However, this relationship was substantially attenuated (and lost precision) following adjustment for preceding, individual-level, sociodemographic and economic determinants of residence in a temporary structure/shack – indicating that poverty was likely to be a far more important barrier to lockdown compliance than informal/temporary housing per se. Nonetheless, the fact remains that two of the personal assets/household services least commonly reported by respondents resident in temporary structures/shacks (a computer and a private/indoor toilet) are also those that are likely to have *directly* undermined these households’ ability to comply with lockdown restrictions by: limiting opportunities for ‘working (virtually) from home’; and requiring residents to leave their home to use the toilet/dispose of human waste.^52-54^ Indeed, a post hoc examination of two items in the AB-R8 survey (in which: all respondents were asked how often they used the internet; and those 1,255/1,381 [90.9%] who personally owned a mobile phone were asked whether these had access to the internet) reveal that those living in temporary structures/shacks: were far less likely to access the internet “Every day” (OR: 0.61; 95%CI: 0.41,0.89); and were also less likely to have access to the internet on their mobile phones (OR: 0.82; 95%CI: 0.53,1.27), when compared to those living in traditional/formal houses/flats/rooms/hostels (although the second of these associations lacked precision; see Table S3.1 and S3.2 in Section 3 of the Supplementary Materials).

These findings are not entirely unanticipated, and given the wealth of research on the social and economic circumstances, and living conditions, of South African households resident in temporary structures/shacks^17,27^ it is frankly astonishing that the constraints these households face were overlooked (and continue to be overlooked) by those responsible for the country’s multi-tiered COVID-19 restrictions.^18,55^ Imposing such restrictions on individuals/households lacking the means to comply not only compounds the existing risks and challenges they face, but also undermines their ability to support and protect themselves (while making them appear responsible for not doing so).^2,52,53^ These residents and communities are not without insight, determination or agency^4,5,7,8,12^ – yet there is little evidence that the South African authorities sought to work with them to develop alternatives to lockdown restrictions that are, at best, impracticable^2,53^ and, at worst, punitive for those who rely on casual, flexible, informal and opportunistic sources of ‘in-person’ (as opposed to ‘virtual’ or ‘online’) work,^2,56^ and those living in contexts where space is at a premium, and where access to clean water, toilet facilities and food necessitate levels of social interaction that require them to breach such restrictions.^18,19,57^

The extraordinary disconnect between what policy makers expect and what shack-dwellers can achieve has led to withering attacks on South Africa’s emergency response to the COVID-19 pandemic.^2,52,53,57,58^ These extend comparable critiques emanating across the globe from analysts and commentators who questioned the feasibility and merits of prolonged lockdowns introduced to reduce the transmission of SARS-CoV2, ‘flatten the curve’ and ensure that health services were not overwhelmed.^48,59,60-63^ Amongst these critics were those who acknowledged the need to impose short-term emergency measures to delay the spread of disease, but who argued that these should only be used to buy the time required to better understand the biology and epidemiology of this new disease; and to better calibrate the costs and benefits of the more extreme and expensive non-pharmacological interventions (NPIs: such as border closures, travel bans, curfews, ‘stay at home’ orders, enforced quarantine and rigorous contact-tracing). While emergency measures were arguably necessary to address the uncertain (and potentially devastating) threat to life posed by a ‘newly emergent disease with pandemic potential’,^64^ by the time most countries across the world were experiencing community transmission of SARS-CoV2, evidence from China^65^ and elsewhere^66^ already provided much needed reassurance that the vast majority of young and healthy people were at low (or very low) risk of ‘serious’ disease (i.e. a level of disease posing a significant threat to life, or warranting professional clinical care). Although there remained extensive uncertainty regarding the transmissibility and longer-term health effects of non-fatal infection^67^ – and even though the promise of effective therapies and vaccines was tainted by academic hubris,^44^ pseudoscience and ‘fake news’^61^ – calls to end lockdown restrictions gained ground and continue to pose a growing challenge to the authority of governments and their scientific advisors.

Amongst these critics were clinicians, epidemiologists and other scholars who questioned the very basis of NPIs on two specific grounds: first, because the much vaunted benefits of these interventions often overlooked or discounted their economic and social costs (including their direct and indirect effects on health and health care);^59-61,68^ and second, because the widespread adoption of universal/blanket (or “one-size-fits-all”)^58^ restrictions failed to recognise the unequal distribution of their potential costs (and likely benefits) to different sectors of the population, and assumed that everyone had access to the resources required to comply).^2,60^ To these we are tempted to add a third, namely that ‘flattening the curve’ to protect finite health services makes little sense (and may even be inequitable) in contexts (or to communities) where these services were already inaccessible or had little to offer *before* the pandemic struck.^20,69,70^ Yet evidence for this proposition (based on data from two further items within the AB-R8 survey) is somewhat equivocal. The first of these required the fieldworker to answer the question *“Are the following facilities present in the primary sampling unit/enumeration area or in easy walking distance?”* – for which one of the facilities listed was *“Health clinic (private or public or both)”*; while the second asked all respondents *“How well or badly would you say the current government is handling the following matters, or haven’t you heard enough to say?”* – for which one of the “matters” included was *“Improving basic health services”* and the five pre-categorised answers were *“Very badly”, “Fairly badly”, “Fairly Well”, “Very well”* and *“Don’t know/Haven’t heard enough”*. Post hoc analyses of these variables reveal that respondents living in temporary structures/shacks were: *less* likely to have been located in primary sampling units/enumeration areas within easy walking distance of a health clinic (OR: 0.71; 95%CI: 0.49,1.03); but somewhat *more* likely to say that improving basic health services had been handled well by the current government (OR: 1.40; 95%CI: 0.96,2.03), when compared to those living in traditional/formal houses/flats/rooms/hostels (although both of these associations lacked precision, and the latter is likely to be substantially confounded by the political affiliation of the respondents involved; see Table S4.1 and S4.2 in Section 4 of the Supplementary Materials).

Hindsight is a cruel teacher, and it is all too easy to forget the uncertainty, anxiety, confusion and wild speculation that accompanied the onset of the COVID-19 pandemic a little over two years ago. Under these circumstances, policy makers might be forgiven for focussing on the risks posed by a potentially devastating new disease, and for overlooking the direct and indirect health effects of lockdown restrictions.^68^ They might also be forgiven for adopting a “one-size-fits-all” approach if only to facilitate public understanding, acceptance and compliance. And no one doubts the pressure they faced to protect health services, particularly where these are a scarce and precious resource. Indeed, some might argue that contemporary criticisms of these decisions can be dismissed on this basis alone, or by pointing out that it is easy to be “wise after the event”. Yet many of these concerns were raised at the very time policy makers were adopting draconian measures while candidly admitting an astonishing degree of uncertainty.^2,57^ And none of these explanations justify the failure of governments across the world to consider the unintended consequences of their decisions, or to apply the established tenets of evidence-informed decision-making in which any intervention (however well-intention or ostensibly beneficial) merits careful scrutiny and monitoring to guard against any known and unknown ill-effects, respectively. Instead, their failure offers perhaps the clearest insight yet into: the hegemony of different sources and forms of knowledge; the dominance of curative medicine over preventative measures/public health; and the limited awareness and understanding of those in authority/power concerning the day to day lives (pressing needs, and legitimate concerns) of ‘ordinary’ people.

Our study sought to focus on the last of these concerns, and while it hopes to have demonstrated the futility of the second, it failed to address the first since it relied upon quantitative techniques that offer more acceptable, manageable and comfortable ‘evidence’ for policy-makers than those generated through qualitative, naturalistic and democratic forms of enquiry. While our analyses nonetheless paint a stark picture of the challenges that households living in temporary structures/shacks face – and the cumulative risks to their health of poverty, under-employment, poor housing and inadequate service provision^2,13^ – a fuller understanding of the lived experiences of these households will require further research to: challenge and stretch the rather limited insights that our data and techniques permit; and add nuance, tone and hue to the rather sketchy quantitative picture these provide. In particular, additional studies will be required to establish *which* of the multifaceted components of South Africa’s lockdown restrictions were least/most challenging for residents of temporary structures/shacks to adopt, and *why* – detail that was unavailable in the items included in the AB-R8 questionnaire.

## Conclusion

There can be little doubt that South African households living in temporary structures/shacks face a number of direct and indirect threats to health, not only as a result of the inadequate protection such dwellings provide (and the additional risks that informal structures and settlements entail), but also as a consequence of the dire social and economic circumstances that forced/led these households to seek shelter therein. These threats to health are compounded whenever ignorance, incompetence, ineptitude or indifference undermine the allocation of finite resources to ensure these households have equitable access to public services and amenities – including regulations intended to safeguard public health during ordinary, and extraordinary, circumstances.

We are not the first to argue that the adoption of a universal/blanket approach to the non-pharmaceutical interventions (NPIs) implemented during South Africa’s response to the COVID-19 pandemic failed to acknowledge the unequal distribution of their impracticability, costs and consequences in one of the world’s most unequal societies. However, our analyses provide robust, quantitative evidence that poverty and inadequate household services are likely to be more important determinants of compliance with NPIs than informal/temporary housing per se. These findings have a number of potential implications for the development of practical, effective and equitable policies that aim to address the collective risks that affect us all, both now and in the future. These implications are particularly important for those risks/policies that impose restrictions on the freedoms we require to survive and thrive – restrictions that can only be justifiable if they are practicable for all, while universal in their benefits, and equitable in their costs and consequences.

- First: universal/blanket policies (such as those intended to be “one-size-fits-all”) can only be equitable when these do not impose impracticable constraints or untenable consequences on those who lack the means to comply (or to prevail thereafter).
- Second: stratified policies (such as those that are “means-tested”) may often be the only way to ensure that those with limited capacity to cope with restrictive regulations are subject to (less draconian) constraints/sanctions or receive additional support to ensure they can comply.
- Third: ongoing research will be required to ensure that policy-makers know both: *which* components of the restrictive regulations available to them might be particularly challenging or impossible for specific households/communities to adopt; and *how* compliance might be strengthened through stratification or the provision of additional services and support.
- Fourth: regardless of the formal evidence available to them, policy-makers and their specialist technical advisers must draw on the insight of communities most likely to be disproportionately affected by, or least able to comply with, any restrictive regulations to ensure they have access to the first-hand, experiential expertise required to assess whether (and how) these communities might be able to comply.
- Fifth: in the absence of formal evidence or experiential expertise, policy-makers might best *assume* that the most disadvantaged members of society will be unable to comply with any regulations that restrict their ability to seek informal/casual/opportunistic sources of work, or access essential goods, services and amenities beyond the confines of their dwellings or underserved communities.

## Data Availability

The source data are available upon request from: https://afrobarometer.org/data

## Acknowledgements

The analyses presented in this paper would not have been possible without the early release of South African data
from Round 8 of Afrobarometer, which will be available for analysis in due course at: http://www.afrobarometer.org/data. Afrobarometer depends on: substantive financial support from a range of external agencies; the diligence of survey teams; and the patience of survey respondents – to all of whom the authors would like to express their sincere gratitude. We are also grateful to Celimpilo Dladla, Mpfariseni Godfrey Mulaudzi, Peter Mekgwe and Tisa Viviers for kindly translating the Title, Abstract and Significance of the main findings into isiZulu, TshiVenda, Setswana and Afrikaans, respectively.

## Supplementary material

**Section 1: Re-categorisation of 26 items selected from within the main questionnaire and supplementary COVID-19 module used in the postponed Afrobarometer survey (Round 8: AB-R8) undertaken across South Africa between 2 May and 10 June 2021**.

### 1.1 Outcomes

*Residence in a Temporary/Non-temporary Structure (dichotomous)* [shack1_dv]

| AB_R8-Q104. In what type of shelter does the respondent live? | Original |  | Re-categorised |  |  |
| --- | --- | --- | --- | --- | --- |
|  | n | % | Category | n | % |
| Non-traditional/Formal house (1) | 1176 | 73.5 | Formal house/flat (0) | 1383 | 86.4 |
| Flat in block of flats (4) | 88 | 5.5 |  |  |  |
| Traditional house/Hut (2) | 119 | 7.4 |  |  |  |
| Temporary structure/shack (3) | 150 | 9.4 | Shack (1) | 150 | 9.4 |
| Single room in a larger dwelling structure or backyard (5) | 48 | 3.0 | Single room/hostel (0) | 53 | 3.3 |
| Hostel in an industrial compound or farming compound (7) | 5 | 0.3 |  |  |  |
| Other (8) | 14 | 0.9 | Other (.) | 14 | 0.9 |

*Ease of compliance with lockdown restrictions (dichotomous)* [comply2_dv]

| AB_R8-COV5 How easy or difficult was it for you and your household to comply with the lockdown or curfew restrictions imposed by the government? | Original - Yes |  | Re-categorised categorical - Yes |  |  |
| --- | --- | --- | --- | --- | --- |
|  | n | % | Category | n | % |
| I/we did not comply (0) | 3 | 0.2 | Difficult or worse (1) | 942 | 58.9 |
| Very difficult (1) | 471 | 29.4 |  |  |  |
| Difficult (2) | 468 | 29.3 |  |  |  |
| Neither easy nor difficult (3) | 122 | 7.6 | Less than difficult (0) | 506 | 31.6 |
| Easy (4) | 242 | 15.1 |  |  |  |
| Very easy (5) | 142 | 8.9 |  |  |  |
| Not applicable (7) | 150 | 9.4 | Not applicable, Refused or Don't know (.) | 152 | 9.5 |
| Don't know (9) | 2 | 0.1 |  |  |  |

### 1.2 Determinants, correlates and consequences (and potential/candidate confounders)

#### 1.2.1 Sociodemographic characteristics

*Respondent Age (polythomous)* [age_dv]

| AB_R8-Q1. How old are you? | Original |  |  | Re-categorised |  |  |
| --- | --- | --- | --- | --- | --- | --- |
|  | Mean | Min | Max | Category | n | % |
| [Continuous in years, as reported] | 39.0 | 18 | 90 | 18-25 (3) | 366 | 22.9 |
|  |  |  |  | 26-35 (2) | 445 | 27.8 |
|  |  |  |  | 36-50 (1) | 453 | 28.3 |
|  |  |  |  | 51-90 (0) | 335 | 20.9 |
|  |  |  |  | Don't know (.) | 1 | 00.1 |

*Respondent Gender (dichotomous)* [gender_dv]

| AB_R8-Q101. Respondent's gender | Original |  | Re-categorised |  |  |
| --- | --- | --- | --- | --- | --- |
|  | n | % | Category | n | % |
| Male (1) | 799 | 49.9 | Male (1) | 799 | 49.9 |
| Female (2) | 801 | 50.1 | Female (0) | 801 | 50.1 |

*Respondent ‘Population Group’ Classification (polytomous)* [race_dv]

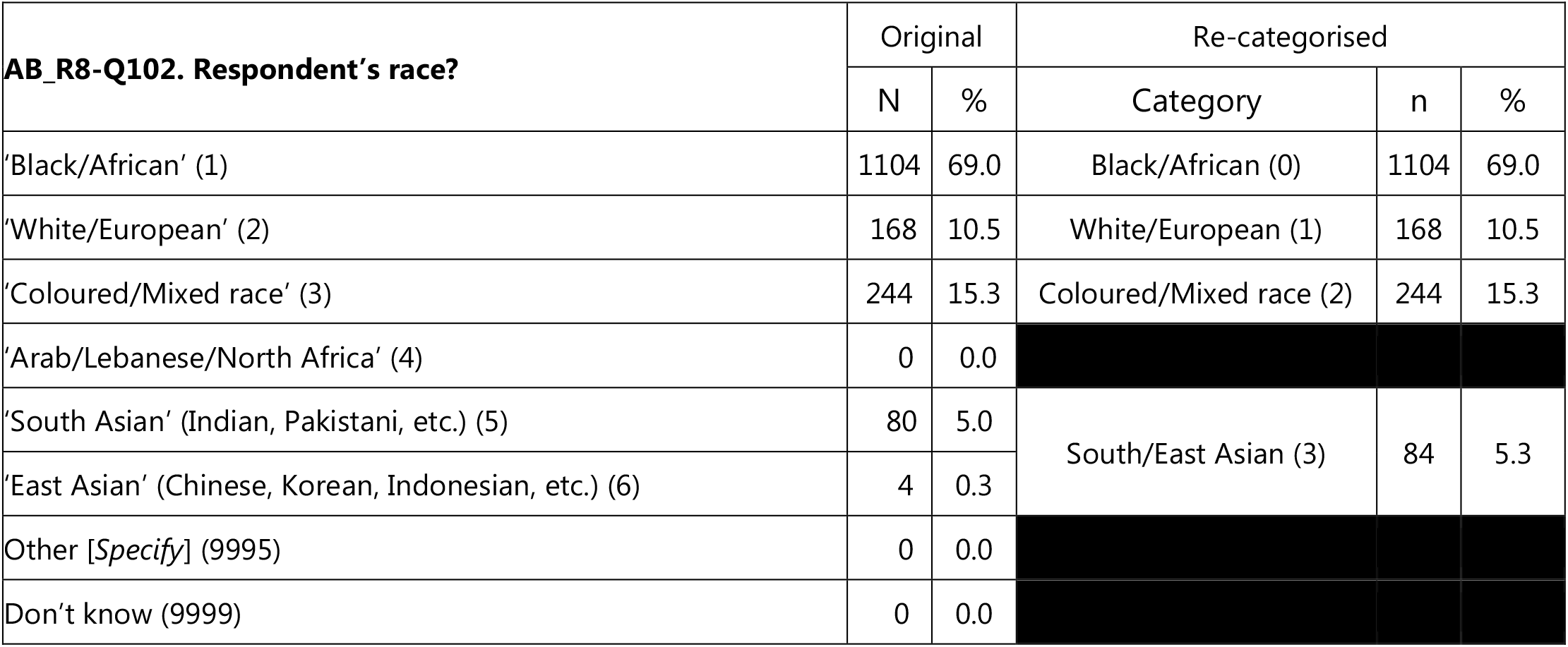

*Respondent Home Language (dichotomous)* [language_dv]

| **AB_R8-Q2. What is the primary language you speak in your home now?** | Original |  | Re-categorised |  |  |
| --- | --- | --- | --- | --- | --- |
|  | N | % | Category | n | % |
| English (1) | 384 | 24.0 | European (1) | 385 | 24.1 |
| French (2) | 0 | 0.0 |  |  |  |
| Portuguese (3) | 1 | 0.1 |  |  |  |
| Swahili (4) | 0 | 0.0 | Non-European (0) | 1215 | 75.9 |
| Other [*Specify*] (various African languages) | 1215 | 75.9 |  |  |  |
| Don't know (9999) | 0 | 0.0 |  |  |  |

#### 1.2.2 Individual-level economic characteristics

*Respondent Educational Attainment (polytomous)* [education_dv]

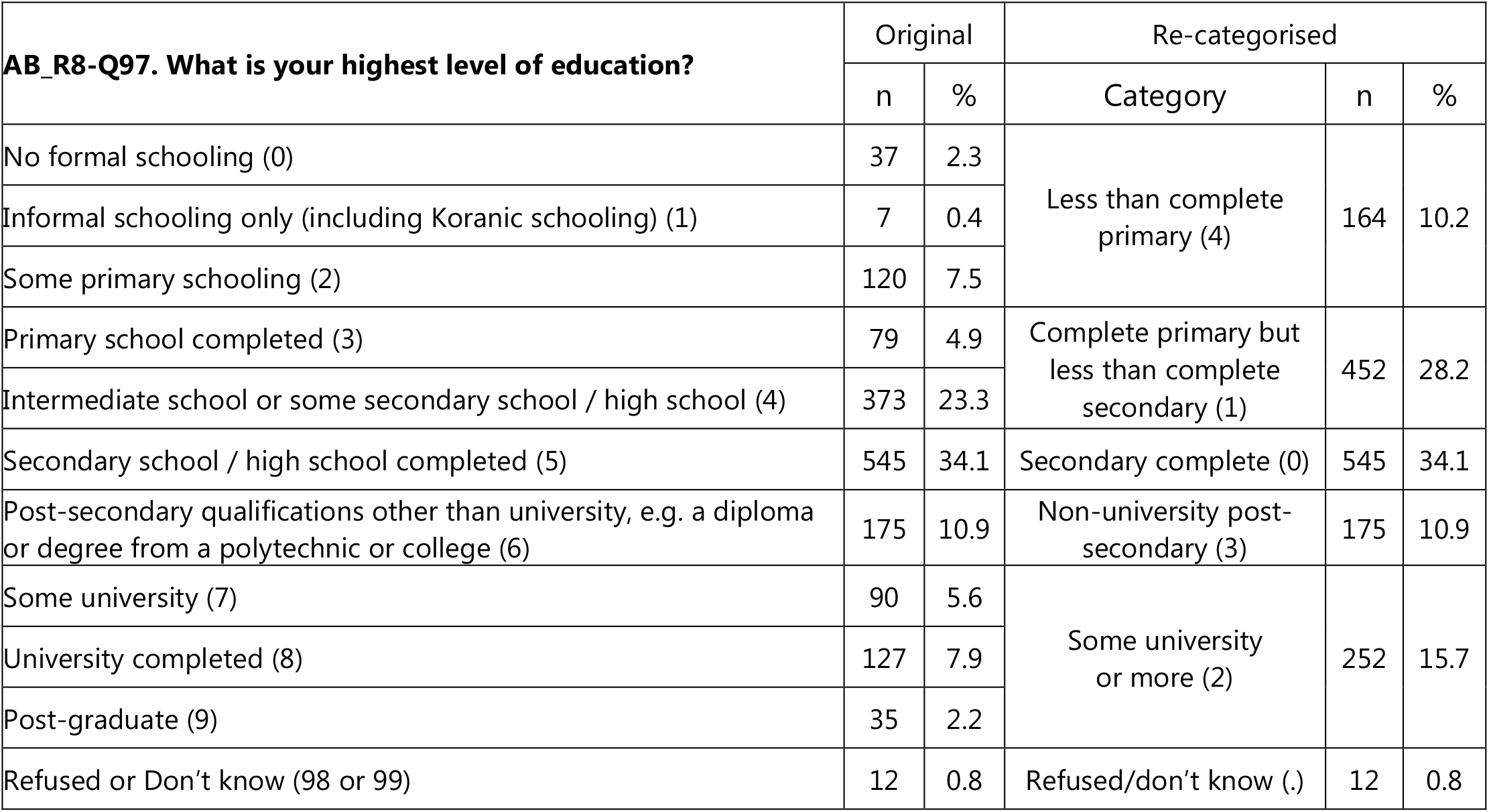

1.2.2 Individual-level economic characteristics *Respondent Educational Attainment (polytomous) [education\_dv]*
| **AB_R8-Q97. What is your highest level of education?** | Original |  | Re-categorised |  |  |
| --- | --- | --- | --- | --- | --- |
|  | n | % | Category | n | % |
| No formal schooling (0) | 37 | 2.3 | Less than complete primary (4) | 164 | 10.2 |
| Informal schooling only (including Koranic schooling) (1) | 7 | 0.4 |  |  |  |
| Some primary schooling (2) | 120 | 7.5 |  |  |  |
| Primary school completed (3) | 79 | 4.9 | Complete primary but less than complete secondary (1) | 452 | 28.2 |
| Intermediate school or some secondary school / high school (4) | 373 | 23.3 |  |  |  |
| Secondary school / high school completed (5) | 545 | 34.1 | Secondary complete (0) | 545 | 34.1 |
| Post-secondary qualifications other than university, e.g. a diploma or degree from a polytechnic or college (6) | 175 | 10.9 | Non-university post-secondary (3) | 175 | 10.9 |
| Some university (7) | 90 | 5.6 | Some university or more (2) | 252 | 15.7 |
| University completed (8) | 127 | 7.9 |  |  |  |
| Post-graduate (9) | 35 | 2.2 |  |  |  |
| Refused or Don't know (98 or 99) | 12 | 0.8 | Refused/don't know (.) | 12 | 0.8 |

*Respondent Employment (polytomous)* [employ_dv]

| **AB_R8-Q95A. Do you have a job that pays a cash income? [If yes, ask:] Is it full time or part time? [If no, ask:] Are you currently looking for a job?** | Original |  | Re-categorised |  |  |
| --- | --- | --- | --- | --- | --- |
|  | n | % | Category | n | % |
| No (not looking) (0) | 381 | 23.8 | No (not looking) (2) | 381 | 23.8 |
| No (looking) (1) | 608 | 38.0 | No (looking) (0) | 608 | 38.0 |
| Yes, part time (2) | 197 | 12.3 | Yes, part time (3) | 197 | 12.3 |
| Yes, full time (3) | 396 | 24.8 | Yes, full time (1) | 396 | 24.8 |
| Refused or Don't know (8 or 9) | 18 | 1.1 | Refused or Don't know (.) | 18 | 1.1 |

*Respondent Assets (dichotomous)* [persradio_dv; perstv_dv; persvehicle_dv; perscomputer_dv; persbank_dv; persmobile_dv]

| **AB_R8-Q92. Which of these things do you personally own...?** | Original - Yes |  | Re-categorised - Yes |  |  |
| --- | --- | --- | --- | --- | --- |
|  | n | % | Category | n | % |
| **92A.** Radio (Yes: 2; No: 1 or 0; Don't know: 9) | 1102 | 68.9 | Radio – Yes (1) | 1102 | 68.9 |
| **92B.** Television (Yes: 2; No: 1 or 0; Don't know: 9) | 1120 | 70.0 | TV – Yes (1) | 1120 | 70.0 |
| **92C.** Motor vehicle or motorcycle (Yes: 2; No: 1 or 0; Don't know: 9) | 453 | 28.3 | Vehicle – Yes (1) | 453 | 28.3 |
| **92D.** Computer (Yes: 2; No: 1 or 0; Don't know: 9) | 582 | 36.4 | Computer – Yes (1) | 582 | 36.4 |
| **92E.** Bank account (Yes: 2; No: 1 or 0; Don't know: 9) | 1305 | 81.6 | Account – Yes (1) | 1305 | 81.6 |
| **92F.** Mobile phone (Yes: 2; No: 1 or 0; Don't know: 9) | 1452 | 90.8 | Mobile – Yes (1) | 1452 | 90.8 |

#### 1.2.3 Household-level economic characteristics

*Dwelling Structure (polytomous)* [shack2_dv]

1.2.3 Household-level economic characteristics *Dwelling Structure (polytomous) [shack2\_dv]*
| **AB_R8-Q104. In what type of shelter does the respondent live?** | Original |  | Re-categorised |  |  |
| --- | --- | --- | --- | --- | --- |
|  | n | % | Category | n | % |
| Non-traditional/Formal house (1) | 1176 | 73.5 | Formal house/flat (0) | 1383 | 86.4 |
| Flat in block of flats (4) | 88 | 5.5 |  |  |  |
| Traditional house/Hut (2) | 119 | 7.4 |  |  |  |
| Temporary structure/shack (3) | 150 | 9.4 | Shack (1) | 150 | 9.4 |
| Single room in a larger dwelling structure or backyard (5) | 48 | 3.0 | Single room/hostel (2) | 53 | 3.3 |
| Hostel in an industrial compound or farming compound (7) | 5 | 0.3 |  |  |  |
| Other (8) | 14 | 0.9 | Other (.) | 14 | 0.9 |

*Household Co-occupancy (polytomous)* [crowding2_dv]

| **AB_R8- ADULT_CT. Total number of adult citizens in the household?** | Original |  | Re-categorised |  |  |
| --- | --- | --- | --- | --- | --- |
|  | n | % | Category | n | % |
| One (1) | 516 | 32.3 | One (0) | 516 | 32.3 |
| Two (2) | 404 | 25.3 | Two (1) | 404 | 25.3 |
| Three (3) | 274 | 17.1 | More than two (2) | 680 | 42.5 |
| Four (4) | 173 | 10.8 |  |  |  |
| Five (5) | 118 | 7.4 |  |  |  |
| Six (6) | 60 | 3.8 |  |  |  |
| Seven to 19 (7-19) | 55 | 3.4 |  |  |  |

*Household assets (dichotomous)* [hhldradio_dv; hhldtv_dv; hhldvehicle_dv; hhldcomputer_dv; hhldbank_dv; hhldmobile_dv]

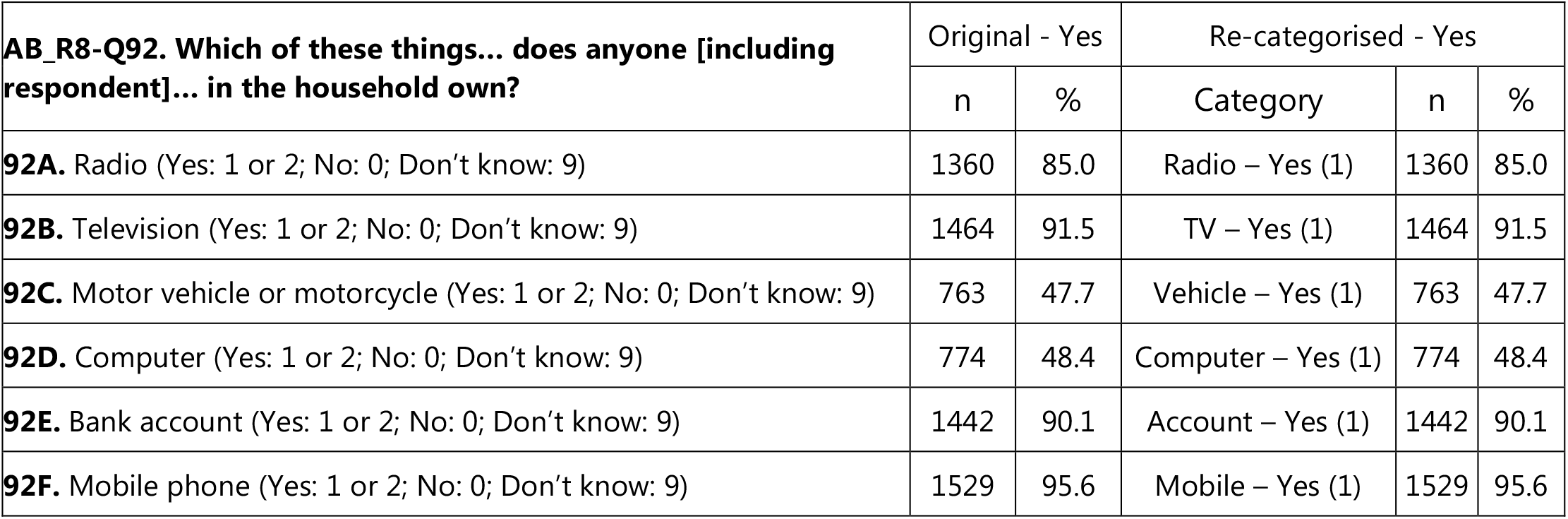

#### 1.2.4 Household services, utilities and amenities

*Household Electricity Supply (dichotomous)* [hhldelectric_dv]

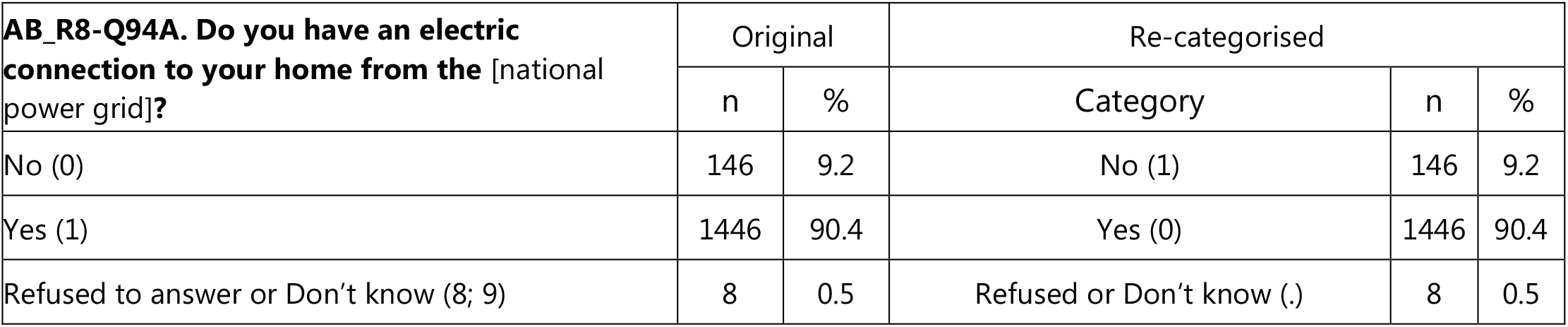

1.2.4 Household services, utilities and amenities *Household Electricity Supply (dichotomous) [hhlelectric\_dv]*
| **AB_R8-Q94A. Do you have an electric connection to your home from the [national power grid]?** | Original |  | Re-categorised |  |  |
| --- | --- | --- | --- | --- | --- |
|  | n | % | Category | n | % |
| No (0) | 146 | 9.2 | No (1) | 146 | 9.2 |
| Yes (1) | 1446 | 90.4 | Yes (0) | 1446 | 90.4 |
| Refused to answer or Don't know (8; 9) | 8 | 0.5 | Refused or Don't know (.) | 8 | 0.5 |

*Household Water Supply (polytomous)* [hhldwaterinout_dv]

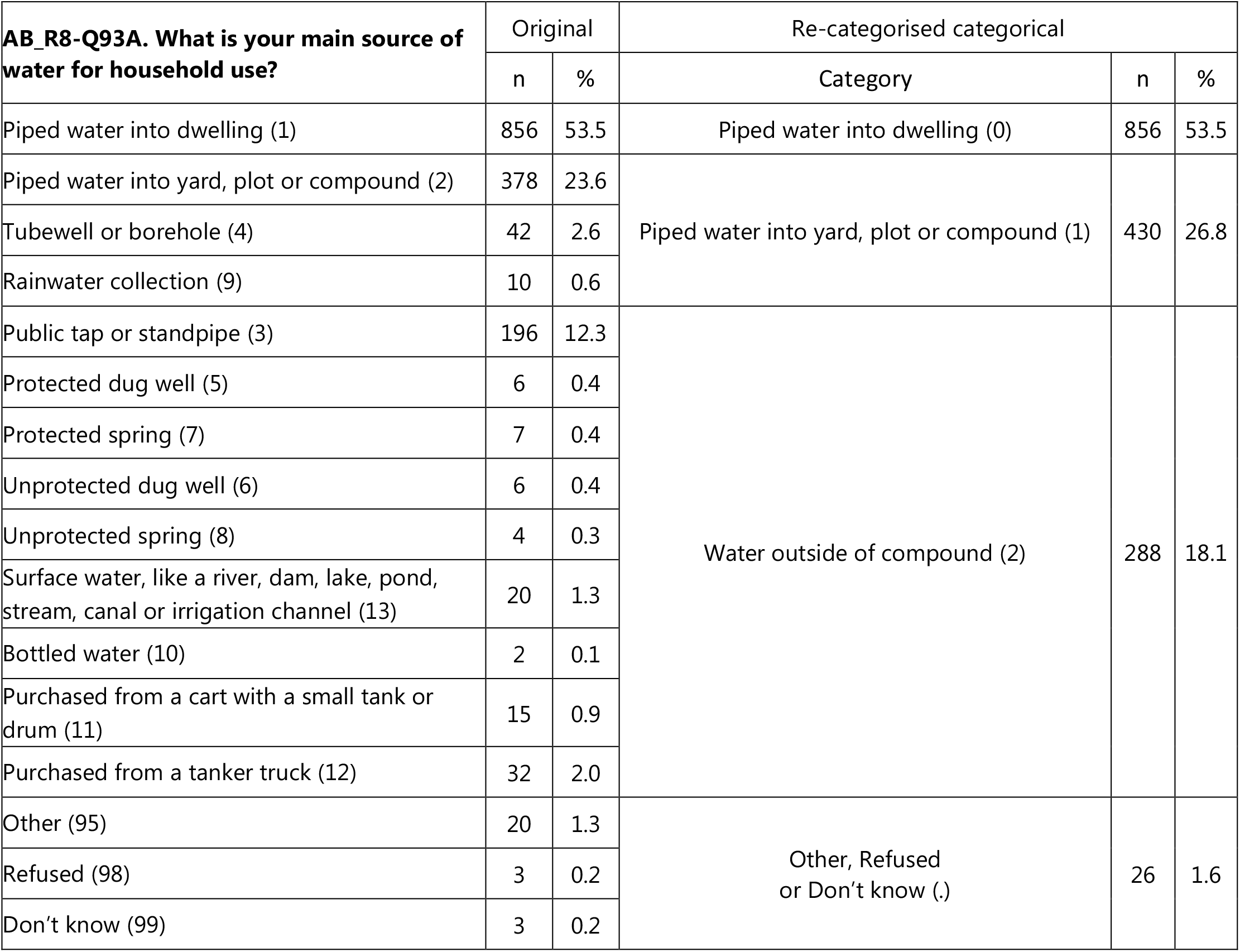

*Household Toilet Facilities (polytomous)* [hhldtoiletinout_dv]

| **AB_R8-Q93B. Do you have a toilet, water closet or latrine available for your use? [If yes] Is it inside your house, inside your compound, or outside your compound, or is there none available?** | Original |  | Re-categorised categorical |  |  |
| --- | --- | --- | --- | --- | --- |
|  | n | % | Category | n | % |
| Yes, inside the house (1) | 810 | 50.6 | Private inside (0) | 810 | 50.6 |
| Yes, inside the compound (2) | 453 | 28.3 | Private inside compound (1) | 453 | 28.3 |
| No, none available (0) | 68 | 4.3 | None or shared/outside compound (2) | 334 | 20.9 |
| Yes, outside the compound (3) | 266 | 16.6 |  |  |  |
| Refused or Don't know (8; 9) | 3 | 0.2 | Refused or Don't know (.) | 3 | 0.2 |

#### 1.2.5 COVID-19 related impacts on health and livelihood

*Household COVID-19 related illness (dichotomous)* [covidill_dv]

1.2.5 COVID-19 related impacts on health and livelihood *Household COVID-19 related illness (dichotomous) [covidill\_dv]*
| **AB_R8-COV2 Please tell me if you personally or any other member of your household have been affected in any of the following ways by the COVID-19 pandemic... Became ill with COVID-19** | Original - Yes |  | Re-categorised categorical – Yes |  |  |
| --- | --- | --- | --- | --- | --- |
|  | n | % | Category | n | % |
| Yes (1) | 284 | 17.8 | Yes (1) | 284 | 17.8 |
| No (2) | 1160 | 72.5 | No (0) | 1160 | 72.5 |
| Not applicable (7) | 150 | 9.4 | Not applicable, Refused or Don't know (.) | 156 | 9.8 |
| Refused (8) | 3 | 0.2 |  |  |  |
| Don't know (9) | 3 | 0.2 |  |  |  |

*Household COVID-19 related job/business/income loss (dichotomous)* [covidjob_dv]

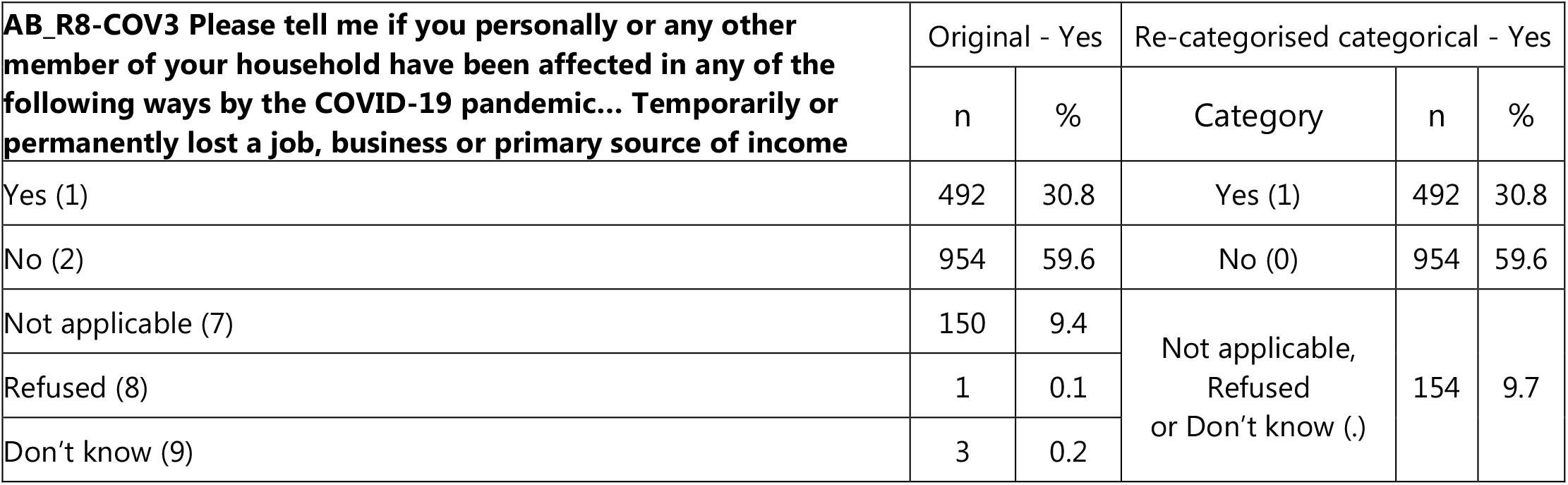

## Section 2: Sensitivity analyses

**Table S2.1.** The sociodemographic and economic determinants of residence in a temporary structure/shack vs. a non-temporary/permanent structure (i.e. a house/flat, room or hostel); and the relationship between residence in a (non)temporary structure/shack and: a number of household-level characteristics (including co-occupancy and household assets, services and amenities); and COVID-19 related illness and/or job/business/income loss – both after adjustment for any preceding potential/candidate confounders (Column S2.1.1 – as in Column 2.2 of Table 2); and after adjustment for all such variables *with the exception of* respondent employment and individual/household assets (Column S2.1.2). All results are presented as odds ratios (ORs) with 95% confidence intervals in parentheses (95%CI).

| **Specified outcome:** |  | Residence in a temporary structure/shack vs. a non-temporary/permanent structure (house/flat, room, hostel) – dichotomous |  |
| --- | --- | --- | --- |
| Outcome referent (0): |  | Non-temporary structure (house/flat, room or hostel) |  |
| Outcome contrast(1): |  | Temporary structure/shack |  |
|  |  | **Covariate adjustment set** |  |
|  |  | **Column S2.1.1** | **Column S2.1.2** |
|  |  | Any preceding covariates<sup>1</sup> | Any preceding covariates except individual/household employment/assets<sup>2</sup> |
| **Specified exposures:** |  | OR (95%CI) | OR (95%CI) |
| **Covariates considered likely to precede (and be potential determinants of) residence in a temporary structure:<sup>3</sup>** |  |  |  |
| **Respondent Age** | 18-25yr (3) | **3.23 (1.60,6.50)** | As for Column S2.1.1 |
|  | 26-35yr (2) | **3.05 (1.54,6.06)** |  |
|  | 36-50yr (1) | **2.76 (1.37,5.55)** |  |
|  | 51-90yr (0) | Referent |  |
| **Respondent Gender** | Female (0) | Referent | As for Column S2.1.1 |
|  | Male (1) | 1.11 (0.77,1.61) |  |
| **Respondent 'Population Group' classification** | 'Black/African' (0) | Referent | As for Column S2.1.1 |
|  | 'White/European' (1) | **0.11 (0.03,0.46)** |  |
|  | 'Coloured/Mixed race' (2) | 0.78 (0.46,1.35) |  |
|  | 'South/East Asian' (3) | 1 (Empty) |  |
| **Respondent Home Language** | Non-European (0) | Referent | As for Column S2.1.1 |
|  | European (1) | 1.09 (0.65,1.82) |  |
| **Respondent Education** | Less than complete primary (4) | **2.01 (1.04,3.88)** | As for Column S2.1.1 |
|  | Less than complete secondary (1) | 1.53 (0.99,2.35) |  |
|  | Secondary complete (0) | Referent |  |
|  | Non-university post-secondary (3) | **0.38 (0.16,0.91)** |  |
|  | Some university of more (2) | **0.24 (0.08,0.67)** |  |
| **Respondent Employment** | No job (looking) (0) | Referent |  |
|  | No (not looking) (2) | 0.66 (0.39,1.12) |  |
|  | Yes (part time) (3) | 0.57 (0.29,1.09) |  |
|  | Yes (full time) (1) | **0.54 (0.30,0.96)** |  |
| **Respondent Radio** | Do not personally own (0) | Referent |  |
|  | Personally own (1) | 0.77 (0.51,1.16) |  |
| **Respondent Television** | Do not personally own (0) | Referent |  |
|  | Personally own (1) | **0.57 (0.38,0.85)** |  |
| **Respondent Motor Vehicle** | Do not personally own (0) | Referent |  |
|  | Personally own (1) | **0.48 (0.25,0.92)** |  |
| **Respondent Computer** | Do not personally own (0) | Referent |  |
|  | Personally own (1) | **0.55 (0.32,0.93)** |  |
| **Respondent Bank Account** | Do not personally own (0) | Referent |  |
|  | Personally own (1) | 1.25 (0.76,2.05) |  |
| **Respondent Mobile Phone** | Do not personally own (0) | Referent |  |
|  | Personally own (1) | **0.43 (0.25,0.76)** |  |
| **Covariates considered likely to be coterminous with (or determined by) residence in a [non-]temporary structure:** |  |  |  |
| **Household Co-occupancy** | One adult in household (0) | Referent | Referent |
|  | Two adults in household (1) | **1.77 (1.09,2.88)** | **1.79 (1.15,2.77)** |
|  | More than two adults in household (2) | **1.87 (1.17,3.00)** | **2.69 (1.80,4.00)** |
| **Household Radio** | No household member owns (0) | Referent |  |
|  | Household member owns (1) | **0.32 (0.16,0.62)** |  |
| **Household Television** | No household member owns (0) | Referent |  |
|  | Household member owns (1) | **0.23 (0.12,0.43)** |  |
| **Household Motor Vehicle** | No household member owns (0) | Referent |  |
|  | Household member owns (1) | **0.31 (0.16,0.58)** |  |
| **Household Computer** | No household member owns (0) | Referent |  |
|  | Household member owns (1) | 0.62 (0.33,1.18) |  |
| **Household Bank Account** | No household member owns (0) | Referent |  |
|  | Household member owns (1) | 0.53 (0.22,1.31) |  |
| **Household Mobile Phone** | No household member owns (0) | Referent |  |
|  | Household member owns (1) | 0.42 (0.15,1.19) |  |
| **Household Electricity Supply** | Connected to grid (0) | Referent | Referent |
|  | Not connected to grid (1) | **11.80 (7.03,19.81)** | **8.22 (5.31,12.72)** |
| **Household Water Supply** | Water inside dwelling (0) | Referent | Referent |
|  | Water inside compound (1) | **2.82 (1.67,4.77)** | **2.84 (1.88,4.30)** |
|  | Water outside compound (2) | **3.58 (2.09,6.13)** | **3.17 (2.03,4.95)** |
| **Household Toilet Access** | Private inside dwelling (0) | Referent | Referent |
|  | Private inside compound (1) | **4.19 (2.22,7.90)** | **4.66 (2.82,7.69)** |
|  | Outside compound (shared/none available) (2) | **5.78 (3.01,11.11)** | **6.03 (3.58,10.15)** |
| **Household Ease of Lockdown Compliance** | Less than "Difficult" (0) | Referent | Referent |
|  | "Difficult" or worse (1) | 0.99 (0.60,1.63) | 0.98 (0.66,1.47) |
| **Household COVID-19 related Illness** | Did not become ill (0) | Referent | Referent |
|  | Became ill (1) | 1.42 (0.75,2.67) | 0.97 (0.58,1.62) |
| **Household COVID-19 related Job/income lost** | No loss (0) | Referent | Referent |
|  | Lost job/business/income (1) | 1.34 (0.83,2.17) | 1.45 (0.99,2.14) |

<sup>1</sup>Column S2.1.1 (as in Column 2.1 of Table 2): Adjustment for any preceding candidate/potential confounding covariates (and not for coterminous covariates in square parentheses – [...]; with the exception of Respondent Age, Gender, 'Population Group' and Home Language), as listed in the following sequence: Respondent Age, Gender, 'Population Group', Home Language, Respondent Education; Respondent Employment; [Respondent Assets – Radio, Television, Motor Vehicle, Computer, Bank Account, Mobile Phone]; Household Occupancy; [Household Assets – Radio, Television, Motor Vehicle, Computer, Bank Account, Mobile Phone]; [Household Services and Amenities – Electricity, Water, Toilet]; [Lockdown Characteristics – Household Ease of Lockdown Compliance, COVID-19 Illness, Job/Business/Income Loss].
<sup>2</sup>Column S2.1.2: As for Column S2.1.1 except that: Household Occupancy; Household Services and Amenities (i.e. Electricity, Water, Toilet); and Lockdown Characteristics (i.e. Household Ease of Lockdown Compliance, COVID-19 Illness, Job/Business/Income Loss) were **not** adjusted for: Respondent Employment; or Respondent/Household Assets (i.e. Radio, Television, Motor Vehicle, Computer, Bank Account, Mobile Phone).
<sup>3</sup>Alternating white/grey shading indicates groups of covariates considered coterminous (i.e. occurring or crystallising at around the same time, given the questions/items used to ascertain these within the AB-R8 survey). Black shading indicates the omission of individual/household markers of socioeconomic status susceptible to change following the onset of the COVID-19 pandemic, and therefore removed (whether as cause, correlate, consequence or potential/candidate confounder) from all of the analyses summarised in Column S2.2.2.

**Table S2.2.**
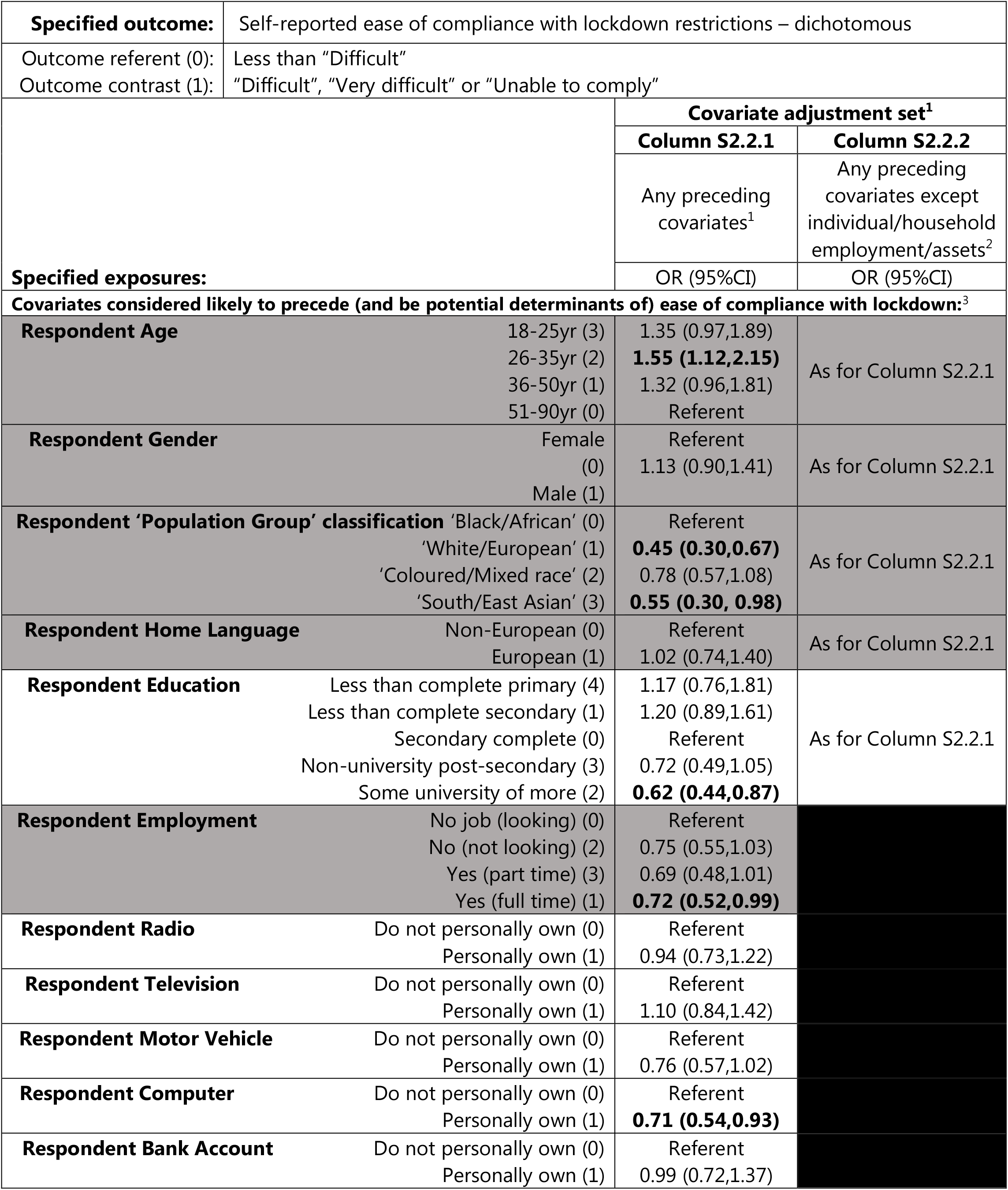
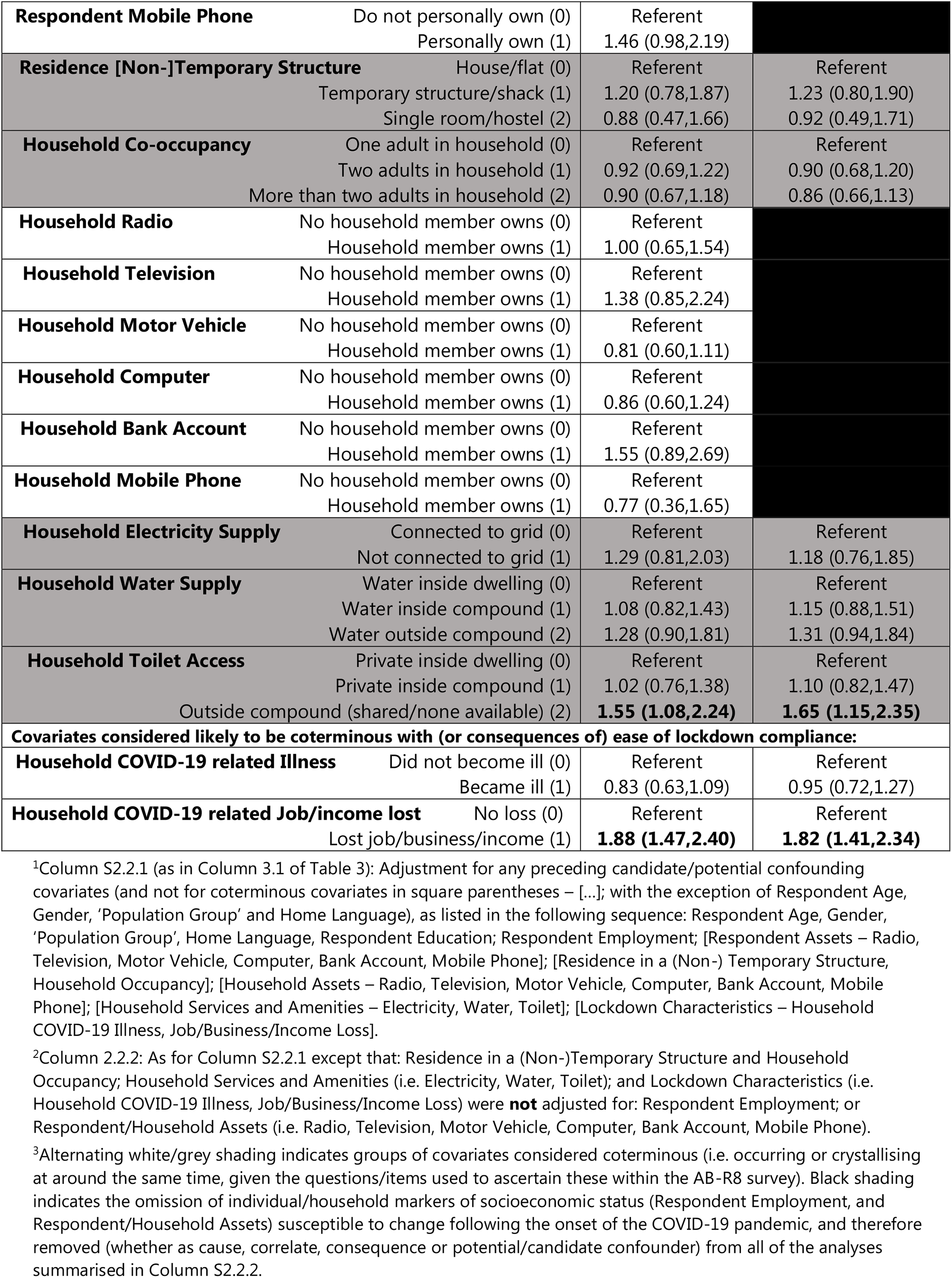
The sociodemographic and economic determinants of ease of compliance with COVID-19 restrictions; and the relationship between ease of compliance and COVID-19 related illness and job/business/income loss; after adjustment for any preceding potential/candidate confounders (Column S2.2.1 – as in Column 3.2 of Table 3); and after adjustment for all such variables with the exception of respondent employment and individual/household assets (Column S2.2.2). All results are presented as odds ratios (ORs) with 95% confidence intervals in parentheses (95%CI).

## Section 3: Post hoc analysis of items within the AB-R8 survey relating to internet access

### 3.1 Access to the internet outcomes

*Frequency of using the internet (dichotomous)* [internetfreq_dv]

| AB_R8-Q92I. How often do you use the internet Internet? | Original |  | Re-categorised |  |  |
| --- | --- | --- | --- | --- | --- |
|  | n | %<br>(n=1,381) | Category | n | %<br>(n=1,381) |
| Never (0) | 368 | 26.6 | Less than every day (0) | 706 | 51.1 |
| Less than once a month (1) | 48 | 3.5 |  |  |  |
| A few times a month (2) | 70 | 5.1 |  |  |  |
| A few times a week (3) | 220 | 15.9 |  |  |  |
| Every day (4) | 649 | 47.0 | Every day (1) | 649 | 47.0 |
| Refused (8) | 3 | 0.2 | Refused/Don't know (.) | 26 | 1.9 |
| Don't know [DNR] (9) | 23 | 1.7 |  |  |  |

*Access to the internet on mobile phone, if own a mobile phone (dichotomous)* [mobileinternet_dv]

| AB_R8-Q92G. Does your phone have access to the Internet? | Original |  | Re-categorised |  |  |
| --- | --- | --- | --- | --- | --- |
|  | N | %<br>(n=1,381) | Category | n | %<br>(n=1,381) |
| No (Does not have Internet access) (0) | 346 | 25.1 | No (0) | 346 | 25.1 |
| Yes (Has Internet access) (1) | 909 | 65.8 | Yes (1) | 909 | 65.8 |
| Not applicable (does not personally have mobile phone) (7) | 118 | 8.5 | Not Applicable/Refused/Don't know (.) | 126 | 9.1 |
| Refused (8) | 1 | 0.1 |  |  |  |
| Don't know [Do not read] (9) | 7 | 0.5 |  |  |  |

**Table S3.1.**
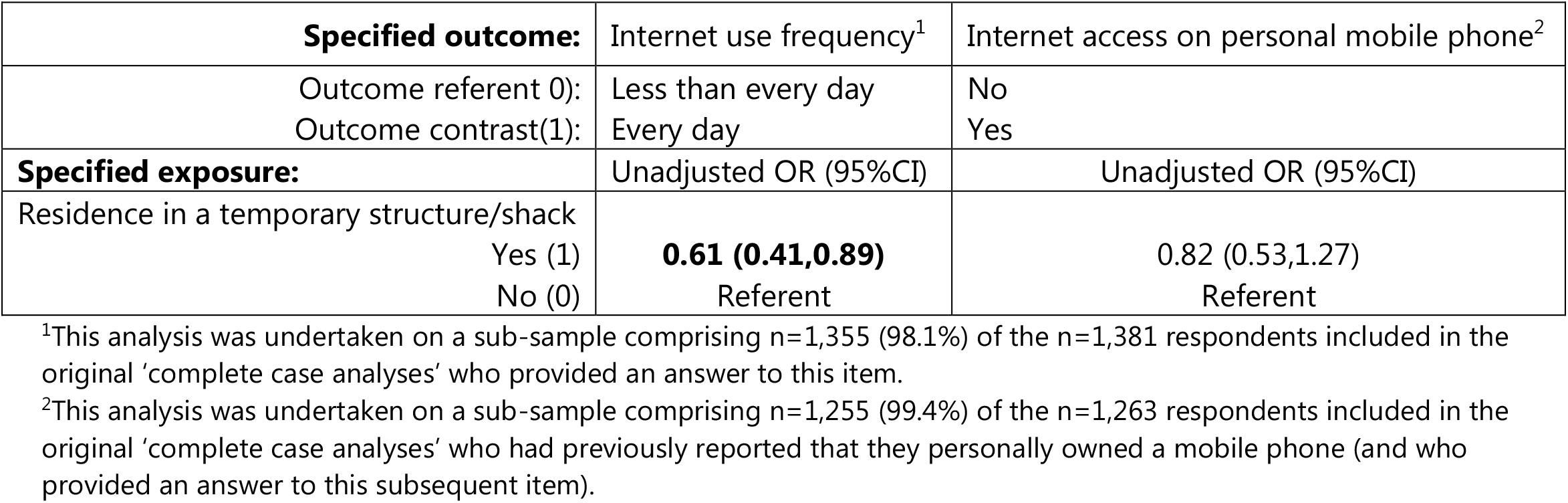
The unadjusted relationship between (non)daily use of the internet, access to the internet on a respondent’s (personal) mobile phone, and residence in a (non)temporary structure/shack. All results are presented as odds ratios (ORs) with 95% confidence intervals in parentheses (95%CI).

## Section 4: Post hoc analysis of items within the AB-R8 survey relating to health care access

### 4.1 Proximity of and government handling, of health services

*Availability of health clinic within sampling unit/enumeration area (dichotomous)* [healthclinic_dv]

| **AB_R8-EA_FAC_D. Are the following facilities [Health clinic (private or public or both)] present in the primary sampling unit/enumeration area or in easy walking distance?** | Original |  | Re-categorised |  |  |
| --- | --- | --- | --- | --- | --- |
|  | n | %<br>(n=1,373) | Category | n | %<br>(n=1,373) |
| Yes (1) | 831 | 60.5 | Yes (1) | 831 | 60.5 |
| No (0) | 513 | 37.4 | No (0) | 513 | 37.4 |
| Can't determine (9) | 29 | 2.1 | Can't determine (.) | 29 | 2.1 |

*Government handling of improving basic health services (dichotomous)* [govthandlehlthwell_dv]

| **AB_R8-Q50 How well or badly would you say the current government is handling the following matters [Improving basic health services], or haven't you heard enough to say?** | Original |  | Re-categorised |  |  |
| --- | --- | --- | --- | --- | --- |
|  | n | %<br>(n=1,381) | Category | n | %<br>(n=1,381) |
| Very badly (1) | 525 | 38.0 | Badly (0) | 760 | 55.0 |
| Fairly badly (2) | 235 | 17.0 |  |  |  |
| Fairly well (3) | 445 | 32.2 | Well (1) | 589 | 42.7 |
| Very well (4) | 144 | 10.4 |  |  |  |
| Refused (8) | 5 | 0.4 | Refused/ Don't know/Haven't heard enough (.) | 32 | 2.3 |
| Don't know/Haven't heard enough [DNR] (9) | 27 | 2.0 |  |  |  |

**Table S4.1.** The unadjusted relationship between the proximity of a nearby health clinic to the sampling unit/enumeration area, respondent views of how the current government is handling improving basic health services, and residence in a (non)temporary structure/shack. All results are presented as odds ratios (ORs) with 95% confidence intervals in parentheses (95%CI).

| **Specified outcome:** | Health clinic nearby<sup>1</sup> | Government handling of health services<sup>2</sup> |
| --- | --- | --- |
| Outcome referent (0): | No | Badly |
| Outcome contrast(1): | Yes | Well |
| **Specified exposure:** | Unadjusted OR (95%CI) | Unadjusted OR (95%CI) |
| Residence in a temporary structure/shack |  |  |
| Yes (1) | 0.71 (0.49,1.03) | 1.40 (0.96,2.03) |
| No (0) | Referent | Referent |

<sup>1</sup>This analysis was undertaken on a sub-sample comprising n=1,352 (97.9%) of the n=1,381 respondents included in the original 'complete case analyses' for whom the fieldworker provided an answer to this item.
<sup>2</sup>This analysis was undertaken on a sub-sample comprising n=1,349 (97.7%) of the n=1,381 respondents included in the original 'complete case analyses' who provided an answer to this item).

